# Intersectional Trajectories in Physical and Mental Health: A UK Longitudinal Study (2009-2024)

**DOI:** 10.64898/2026.10.14.26365511

**Authors:** Daniel Holman, Andrew Bell, Matthew Bennett, Helena Mendes Constante, Mark Green, Aneta Piekut

## Abstract

Health inequalities are often examined through single axes such as class or ethnicity, obscuring how these dimensions interlock. This study applies a longitudinal intersectional approach to examine physical and mental health trajectories across five generations in the UK. We analysed 14 waves of the UK Household Longitudinal Study (2009–2024) using longitudinal Multilevel Analysis of Individual Heterogeneity and Discriminatory Accuracy (MAIHDA). We modelled SF-12 physical and mental health trajectories for 398 intersectional strata defined by gender, ethnicity, occupational class, and generation. Physical health inequalities widened with age, consistent with cumulative disadvantage, while mental health inequalities were more evident among later-born cohorts. The ‘Boomer’ generation exhibited a relatively flat mental health trajectory across its observed age range. In contrast, ‘Generation Z’ had lower predicted mental health, particularly among White British women with no work history – a group with poorer predicted trajectories than their South Asian peers. Our study shows how health inequalities are structurally differentiated by cohort and intersectional position. Policy responses should support proportionate universalism, combining broad population-level support with intensified action for groups showing poorer health.

## Introduction

Socioeconomic position, gender, and ethnic inequalities in physical and mental health are well documented in the UK and internationally (Beckfield et al., 2013; Marmot et al., 2020). Marmot et al.’s (2020) work on health inequalities shows huge variation across English neighbourhoods in life expectancy and disability. Socioeconomic gradients in health operate through both material mechanisms – such as differential access to income and housing – and psychosocial pathways involving stress, discrimination, and perceived control (Blane et al., 1997; Pickett & Wilkinson, 2009). Behavioural factors, including diet, smoking, alcohol consumption, and physical activity, also contribute to the social patterning of health, and are themselves shaped by material and psychosocial conditions (Petrovic et al., 2018).

Life course theory understands lives as unfolding through age-graded trajectories that are shaped by earlier experiences, accumulated advantage and disadvantage, and the historical contexts in which people live (Elder & Giele, 2009). In social epidemiology, this perspective emphasises that health differences between social groups are not fixed at a single point in time, but unfold across age-based trajectories, with inequalities dynamically changing, or persisting with age (Ben-Shlomo et al., 2016; Jones et al., 2019). Inequalities in physical and mental health often emerge early in life and may persist through adulthood and across generations (Hossin et al., 2020; Houweling & Grünberger, 2024; Pearce et al., 2019). Mental health disadvantage from adolescence, for example, is associated with adverse socioemotional development (Black et al., 2023) through to elevated risks of depression and cognitive decline in later life In adulthood and later life (Daly & Allen, 2016; NHS England, 2025). In adulthood and later life, socioeconomic position is associated with transitions to multimorbidity, frailty and disability (Dugravot et al., 2019; Khanolkar et al., 2021; Zaninotto et al., 2020).

Regarding gender, women live longer than men but spend more years in poor health and with disability, reflecting a “male–female health–survival paradox” (Alberts et al., 2014). Across Europe, England and the USA, women report higher levels of disability, arthritis, and depression; whereas men have higher rates of heart disease and mortality (Crimmins et al., 2011). Women tend to report more internalising disorders such as depression and anxiety (NHS England, 2025; Riecher-Rössler, 2017), while men show more externalising behaviours and have higher suicide rates (Boyd et al., 2015). Gender differences in health reflect intertwined social, economic, and biological factors. Gendered social roles shape health-seeking, support, and responses to illness, often exposing women to greater role strain and internalised distress over time (Vlassoff, 2007).

Health inequalities are also structured by ethnicity, though not always in uniformly disadvantageous ways. ONS estimates show striking ethnic and gender differences in life expectancy in England and Wales between 2011 and 2014. White people had the lowest life expectancy (83.1 years for women and 79.7 years for men), while Black African and Bangladeshi groups had among the highest (88.9 and 87.3 years for women, respectively) (Office for National Statistics, 2021). Despite this, accounting for SES, ethnically minoritised groups tend to report poorer self-rated health and more limiting illness than the White British majority (Evandrou et al., 2016; Stopforth et al., 2021). These inequalities persist over time and often widen with age, consistent with cumulative disadvantage linked to socioeconomic inequality, racism, and discrimination (Stopforth et al., 2021). Williams *et al*. (2020) found that South Asian men and women experience elevated functional limitations from midlife onwards, not fully explained by socioeconomic status or comorbidities, while African Caribbean men have lower odds in younger adulthood and African Caribbean women higher risks in later life. Mental health also varies by ethnicity and life stage: Pakistani, Bangladeshi, and Black Caribbean people report poorer mental health than White British counterparts, with socioeconomic disadvantage and racial discrimination identified as important contributors (Wallace et al., 2016). However, a recent UK review found that ethnically minoritised children generally have similar or lower rates of common mental health problems than White peers, though risks vary by group and type of problem (Guan et al., 2024). This evidence demonstrates a complex and nuanced relationship between ethnicity and health.

Although quantitative intersectionality-informed health research has grown, UK population health research often still examines SEP, gender and ethnicity separately, limiting understanding of how they intersect to shape unique positions of advantage and disadvantage. Intersectionality offers a theoretical and methodological framework to address these limitations. Originating in Black Feminist thought, intersectionality theory (Bauer, 2014; Bowleg, 2012) emphasises that social divisions are mutually constitutive rather than additive, operating through interlocking systems of oppression. Following McCall’s (2005) typology, this study adopts an intercategorical approach, using social categories as proxies for social positions within systems of power rather than as causal attributes of individuals. Gender, ethnicity and occupational class are therefore interpreted as markers of unequal social, economic and institutional conditions that may shape health trajectories, while generation captures historically situated context. Applying this framework to quantitative data offers a means of uncovering hidden or counterintuitive patterns of inequality that single-axis approaches can obscure (Evans et al., 2018; Holman & Walker, 2020).

Recent scholarship has brought intersectionality into dialogue with life course epidemiology to examine how inequalities are patterned across age, time, and generation (Ferrer et al., 2017; Holman & Walker, 2020). Synthesising intersectionality with life course analysis is useful because it describes how intersecting social positions shape cumulative advantage and disadvantage across age-based trajectories in historical context. This approach highlights that inequalities may widen, narrow, or persist with age as cumulative exposures interact with social identities and structural positions. However, much intersectionality research remains cross-sectional, limiting understanding of how intersectional inequalities vary across age-related health trajectories, despite evidence that cumulative and intersecting disadvantages may shape later-life health in distinctive ways (Ben-Shlomo & Kuh, 2002; Calasanti & King, 2015; Kuh et al., 2025; Richardson & Brown, 2016).

Cohort or ‘generational context’ reflects the life course principle of historical time and place, which emphasises that lives unfold within particular social, economic, cultural and policy contexts (Elder & Giele, 2009), and is increasingly recognised as an important dimension of health inequality (Gimeno et al., 2025; Hossin et al., 2020; Konstantinou & Attia, 2023). These contexts may shape material conditions, health behaviours, and psychosocial resources differently across cohorts (Gilleard, 2004). Cohorts may therefore be differentially affected because the timing of exposure matters: the same social, economic, or policy change may have different implications depending on whether it is encountered in childhood, early adulthood, midlife, or later life.

For physical health, these cohort differences may reflect changing patterns of work, diet, environmental exposures, and medical and technological advances. International analyses suggest ‘generational drifts’ in physical health in some contexts, whereby later-born cohorts exhibit poorer health at equivalent ages (Gimeno et al., 2024). For mental health, shifting social norms, economic precarity, and digital connectivity can alter stress, identity, and social support structures (Bell et al., 2024; Konstantinou & Attia, 2023). National data show that the proportion of young adults with a common mental health condition increased from 17.5% to 25.8% between 2007 and 2023/4 (NHS England, 2025). Generational differences may therefore be associated with divergent health trajectories: physical health may reflect changing material, occupational, behavioural, and medical contexts, while mental health may be especially sensitive to shifting economic, psychosocial, and digital factors. Integrating generation into a longitudinal intersectional framework allows these historically situated inequalities to be examined.

The present study makes a novel contribution by applying longitudinal MAIHDA modelling to examine physical and mental health trajectories across generations in the UK. We draw on the life course concepts of trajectories and cumulative disadvantage to examine whether health inequalities widen, narrow, or persist across observed age ranges, and whether these patterns vary by intersectional position. Generation is incorporated as both an intersectional factor and a marker of historically situated context, recognising that cohorts age through different social, economic, and policy environments. Rather than deriving specific hypotheses, the analysis directs attention to whether health inequalities unfold differently across intersectional positions and generations, consistent with the use of MAIHDA as an exploratory tool for describing complex patterns of heterogeneity. In mapping how physical and mental health trajectories vary across intersectional positions and generations, we aim to add to the growing body of quantitative intersectionality research and provide evidence relevant to more nuanced approaches to health inequalities.

## Methods

### Data

We analysed data from 14 waves (2009-2024) of the nationally representative UK Household Longitudinal Study (UKHLS). The UKHLS provides a representative longitudinal sample of the UK population using a clustered and stratified probability design, with an ethnic minority boost sample. Our analytic sample included 83,304 individuals aged 16–103 covering 483,887 observations.

Outcomes were the SF-12 Physical Component Summary score (SF-12 PCS) and Mental Component Summary score (SF-12 MCS), scored using the standard SF-12 approach (Ware et al., 1996). This scoring approach has been cross-validated internationally, including in the UK (Gandek et al., 1998), and is used here to aid interpretation of relative differences within the analytical sample. The SF-12 PCS reflects how well a person feels they can carry out their physical activities, and the SF-12 MCS reflects their emotional well-being and how mental health affects daily life. Differences of around 5-10 points are clinically meaningful: a 5-point gap often distinguishes those with mild versus severe chronic conditions, while a 10-point gap is comparable to the burden of serious illness (e.g., congestive heart failure) or clinical depression (Ware et al., 1995). Both measures are widely used for capturing socio-demographic and longitudinal variations in health (Parra-Mujica et al., 2021; Reed et al., 2025).

Intersectional strata were defined based on gender (men, women), ethnicity (White British, White Irish, Other White, Indian, Pakistani, Bangladeshi, Caribbean, African, Mixed, Other), socioeconomic position (managerial and professional, intermediate, routine and manual, no work history) and generation (Silent, Boomer, X, Y, Z) (see below for further details). There were 398 strata in total with a median size of 194 observations and a range of 1 to 25,283. This specification balanced parsimony with intersectional granularity, avoiding further aggregation of social groups while relying on MAIHDA shrinkage to stabilise estimates for smaller strata. Full stratum-level counts are provided in Supplementary Data S1.

While the UKHLS records binary biological sex, we use the term ‘gender’ throughout this study to align with our intersectional framework. Due to these data limitations, this was measured as a binary variable, and individuals whose responses changed across waves (n=113; 0.12%) were excluded. To uncover granular ethnic differences while maintaining sufficient sample sizes for intersectional strata, ethnicity was measured using a 10-category version of the UKHLS 18-category variable (McFall et al., 2024). We retained major ethnic identities individually but consolidated four specific mixed-heritage backgrounds into a single ‘Mixed’ category and grouped smaller, diverse populations (e.g., Arab, Chinese, and Gypsy or Irish Traveller) into a combined ‘Other’ category (see Table 1).

**Table 1.** Analytical sample.

| Variable Name |  |  |
| --- | --- | --- |
| Individual Level (n=83,304) |  | n (%) |
| Ethnicity | White British | 63,210 (75.88%) |
|  | African | 2148 (2.58%) |
|  | Bangladeshi | 1684 (2.02%) |
|  | Caribbean | 1697 (2.04%) |
|  | Indian | 3285 (3.94%) |
|  | Mixed | 1633 (1.96%) |
|  | Other | 2370 (2.85%) |
|  | Other White | 3025 (3.63%) |
|  | Pakistani | 2832 (3.40%) |
|  | White Irish | 1420 (1.70%) |
| Generation | Silent | 12,870 (15.45%) |
|  | Boomer | 22,684 (27.23%) |
|  | X | 21,604 (25.93%) |
|  | Y | 19,937 (23.93%) |
|  | Z | 6209 (7.45%) |
| NS-SEC | Managerial and professional | 25,952 (31.15%) |
|  | Intermediate | 17,691 (21.24%) |
|  | Routine and manual | 31,997 (38.41%) |
|  | No work history | 7664 (9.20%) |
| Gender | Male | 38,091 (45.73%) |
|  | Female | 45,213 (54.27%) |
| Observation Level (n=483,887) |  | Mean/SD |
| Age |  | 49.41 (18.11) |
| SF-12 Physical Component Summary score |  | 49.34 (11.19) |
| SF-12 Mental Component Summary score |  | 48.86 (10.34) |

NS-SEC = National Statistics Socio-Economic Classification

Socioeconomic position was measured using occupational class (National Statistics Socio-Economic Classification (NS-SEC)). Analysing occupational class is more consistent with intersectional theory than using education or income, since it is a group-based measure that captures relationality, identity, and discrimination, in line with other conventional axes of inequality (Bauer, 2014; Rose & Pevalin, 2003). At the observation level, a composite three-category NS-SEC was created that prioritised current job NS-SEC where available, then last job, and finally first job (managerial and professional, intermediate, routine and manual). Valid responses were retained, while missing codes were not. At the person level, a modal NS-SEC class was assigned; in cases of ties, the most recent valid wave was used. This person-level measure captures dominant observed occupational class location across the study period, rather than a time-varying exposure or strictly prior measure of SEP; sensitivity analyses using first observed valid NS-SEC produced substantively similar findings (Supplementary Tables S1 and S2). Job status was used when no valid NS-SEC was available to determine the reason: individuals never observed as employed or self-employed were coded ‘no work history’. We set NS-SEC to missing only where there was no evidence of work and all available NS-SEC entries across waves were missing. This is a pragmatic solution to the fact that NS-SEC is based on employment relations and conditions, and cannot be straightforwardly applied to those outside paid work. Treating this group as analytically distinct avoids excluding them altogether which is important given that economic inactivity is socially patterned and associated with poorer health outcomes (Crawshaw et al., 2024).

Generations were categorised by birth year following the groupings used by Bell et al. (2024): Silent (≤1945), Boomer Generation (1946–1963), Generation X (1964–1979), Generation Y (Millennials; 1980–1994), and Generation Z (1995 onwards). Generational categories are heuristic, tied to specific cultural contexts, rather than fixed: different sources define boundaries differently, and individuals born near the cut-offs may plausibly be considered members of two generations. Our analysis therefore treats generations as approximate birth-year bands, acknowledging that these boundaries are socially constructed and contested rather than natural divisions (Aboim & Vasconcelos, 2014; Rudolph & Zacher, 2022). Predicted differences between generations should therefore be interpreted cautiously, particularly for individuals close to generational cut-offs. Supplementary Table S3 reports the observed age distribution by generation. Generational contrasts are interpreted descriptively, particularly where non-adjacent generations have little or no age overlap.

### Missing data

The SF-12 physical and mental health scales are derived from items included in the self-completion questionnaire. Observations where individuals did not complete the questionnaire (6.41%, n=36,484) or were proxy respondents (5.17%, n=29,404), or had missing outcome data (1.12%, n=6,360) were ineligible and excluded. A further 12,811 observations were removed because individuals were in education across all waves, had inconsistent or missing demographic data (generation, gender, or ethnicity), or were under 16. The final analytical sample comprised 483,887 observations across 83,304 individuals. Individuals contributed a median of 4 observations (range 1–14; Supplementary Table S4); across the 398 intersectional strata, the median stratum contained 45 individuals.

### Analysis

We used the longitudinal extension to MAIHDA developed by Bell et al. (2024). Cross-sectional MAIHDA models use multilevel modelling to nest individuals (level 1) within intersectional strata (level 2), defined by combinations of social characteristics (Evans et al., 2024). The model estimates the extent to which variation in an outcome is partitioned between individuals and intersectional strata, revealing the extent of intersectional clustering. This provides more precise estimates of intersectional differences than conventional approaches using interaction effects or subgroup analysis through shrinkage (Bell et al., 2019).

Longitudinal MAIHDA extends this approach by nesting observations within individuals within intersections, in a three-level model (see Figure 1). This approach provides estimations of age-related trajectories in health to differ between intersectional strata and enables comparison of the proportion of heterogeneity captured at the individual vs intersectional level.

**Figure 1.**
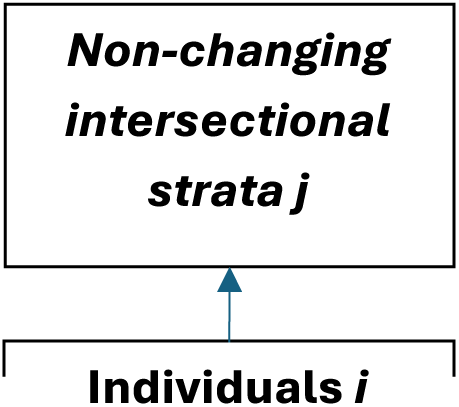
Longitudinal MAIHDA Structure.

Two main models are specified. In Model 1, age (mean-centred) is included as a time-varying covariate in the fixed part of the model as a cubic polynomial to capture age-related health trajectories, following Bell et al. (2024) and supported by AIC comparisons with simpler age specifications (Supplementary Tables S5 and S6). In contrast to cross-sectional models, random-slopes associated with age are included. Only the linear age term was allowed to vary at the individual and stratum levels; quadratic and cubic age terms were included as fixed effects. We estimate an overall age trajectory for health, while also allowing both intersectional strata and individuals to deviate from that average pattern. At the stratum level, we capture variability not only in the average level of health but also in the rate of age-related change, reflecting differences in health trajectories across intersectional positions. At the individual level, the model allows each person to have their own starting point and rate of age-related change, rather than assuming everyone in the same stratum follows an identical trajectory. This recognises that even among people who share the same intersectional position, there is additional heterogeneity in how health develops over time. Finally, at the occasion level, we capture within-individual fluctuations in health across repeated measurements. Taken together, this structure enables us to model age-related trajectories as both socially patterned and individually variable, while accounting for change over time.

Model 2 then builds on model 1, adding in both the main effects for the stratum-defining variables, and interactions between those variables and the linear component of age. The age-by-generation interactions therefore indicate whether the linear age slope differs between each generation and the Silent generation reference group, conditional on the shared fixed quadratic and cubic age terms. This results in better strata trajectory predictions (Leckie et al., 2025) whilst also revealing the extent of multiplicativity in both the intercepts and age slopes. Because household and other contextual clustering were not modelled, any stable variation associated with these contexts may be partly absorbed into the individual-level variance component. The model accounts for stratum-level, individual-level and within-individual variability, but does not include additional contextual levels such as households, geographical areas or workplaces. Including these structures was not computationally feasible in the current longitudinal MAIHDA framework. Stable variability associated with these contexts may therefore be partly absorbed into the individual-level variance component. Further technical details on the modelling approach are included in the Supplementary material, including the interpretation of intercept and slope VPCs, PCVs and age-varying variance functions.

To help visualise the results we developed an RShiny app, based on visualisation in Bell et al. (2024), available at: https://inequalities-research.shinyapps.io/IntersectExplorer/

Further details including technical documentation are available on GitHub: https://github.com/danjholman/IntersectExplorer-Shiny.

## Results

Descriptive statistics for the analytical sample are presented in Table 1.

The main results are presented in Table 2 (full results are given in Supplementary Tables S5 and S6). Model 1P shows that physical health declines with age, with statistically significant squared and cubic terms indicating modest non-linearity in an overall declining age trajectory. The random effects indicate substantial between-individual differences in mean SF-12 PCS: ∼18% of which is attributable to intersectional strata. Variation in age-related change is small, with age slope variances of 0.008 at level 3 and 0.042 at level 2, indicating little heterogeneity in rates of decline. These values suggest that inequalities in physical health are driven more by differences in centred-age health than by rates of decline.

**Table 2.** Results from Longitudinal MAIHDA Models Predicting Physical and Mental Health Trajectories.

|  | Model 1P, PCS |  | Model 2P, PCS |  |  |  | Model 1M, MCS |  | Model 2M, MCS |  |  |  |
| --- | --- | --- | --- | --- | --- | --- | --- | --- | --- | --- | --- | --- |
|  | Estimates | SE | Estimates | SE | Estimates | SE | Estimates | SE | Estimates | SE | Estimates | SE |
| Intercept | 48.269*** | .215 | 55.156*** | .552 |  |  | 45.604*** | .273 | 55.713*** | .620 |  |  |
| Age (mean-centred) | -.274*** | .008 | -.215*** | .027 |  |  | -.033*** | .010 | -.211*** | .027 |  |  |
| Age squared | -.00437*** | .00016 | -.00513*** | .00026 |  |  | .00328*** | .00021 | .00346*** | .00030 |  |  |
| Age cubed | -.00003*** | .00000 | -.00002*** | .00001 |  |  | -.00009*** | .00001 | -.00009*** | .00001 |  |  |
|  |  |  | Main effects |  | Age interactions |  |  |  | Main effects |  | Age interactions |  |
| African (ref White British) |  |  | -0.768 | .486 | -.034 | .022 |  |  | 2.604*** | .551 | .017 | .023 |
| Bangladeshi |  |  | -3.941*** | .524 | -.051* | .024 |  |  | -.092 | .586 | -.039 | .024 |
| Caribbean |  |  | -1.035* | .484 | -.030 | .022 |  |  | 1.197* | .556 | .062** | .023 |
| Indian |  |  | -1.495*** | .444 | .002 | .020 |  |  | 1.375** | .518 | -.026 | .020 |
| Mixed |  |  | -0.524 | .509 | -.005 | .023 |  |  | -1.027 | .569 | .006 | .023 |
| Other |  |  | -1.221** | .463 | -.032 | .021 |  |  | .483 | .532 | -.004 | .021 |
| Other White |  |  | 0.734 | .455 | .035 | .020 |  |  | 1.716** | .527 | .024 | .021 |
| Pakistani |  |  | -3.465*** | .465 | -.040 | .021 |  |  | .121 | .533 | -.021 | .021 |
| White Irish |  |  | -0.138 | .484 | .009 | .021 |  |  | 1.408* | .550 | .017 | .022 |
| Boomer (ref Silent Gen) |  |  | -3.243*** | .519 | .085*** | .024 |  |  | -6.172*** | .545 | .133*** | .023 |
| Gen X |  |  | -2.507*** | .526 | -.040 | .027 |  |  | -8.110*** | .558 | .088** | .028 |
| Gen Y |  |  | -3.027*** | .527 | -.070* | .032 |  |  | -11.540*** | .630 | .098*** | .034 |
| Gen Z |  |  | -1.444 | .861 | -.027 | .040 |  |  | -18.766*** | .996 | .015 | .044 |
| NS-SEC Intermed. (ref Manag. & pro.) |  |  | -1.498*** | .319 | -.001 | .015 |  |  | -.361 | .361 | -.001 | .015 |
| Routine and manual |  |  | -3.475*** | .303 | -.044** | .014 |  |  | -1.162*** | .346 | .001 | .014 |
| No work history |  |  | -7.868*** | .383 | -.096*** | .017 |  |  | -4.514*** | .408 | -.042** | .016 |
| Female (ref Male) |  |  | -1.369*** | .233 | -.037*** | .011 |  |  | -1.403*** | .266 | .043*** | .011 |
| **Random effects** |  |  |  |  |  |  |  |  |  |  |  |  |
| Level 3 intercept variance | 12.862 |  | 2.222 |  |  |  | 22.277 |  | 3.493 |  |  |  |
| Level 3 age-slope variance | .008 |  | .004 |  |  |  | .009 |  | .004 |  |  |  |
| Level 3 intercept-slope covariance | .152 |  | .032 |  |  |  | .091 |  | .074 |  |  |  |
| Level 2 intercept var | 56.868 |  | 56.870 |  |  |  | 46.233 |  | 46.351 |  |  |  |
| Level 2 age-slope variance | .042 |  | .042 |  |  |  | .019 |  | .019 |  |  |  |
| Level 2 intercept-slope covariance | 1.201 |  | 1.201 |  |  |  | .022 |  | .020 |  |  |  |
| Level 1 variance | 37.945 |  | 37.945 |  |  |  | 52.077 |  | 52.080 |  |  |  |
| Slope VPC: L3/(L2+L3) | 15.37% |  | 9.15% |  |  |  | 30.95% |  | 17.44% |  |  |  |
| Intercept VPC: L3/(L2+L3) | 18.45% |  | 3.76% |  |  |  | 32.52% |  | 7.01% |  |  |  |
| Slope PCV |  |  | 44.48% |  |  |  |  |  | 53.62% |  |  |  |
| Intercept PCV |  |  | 82.73% |  |  |  |  |  | 84.32% |  |  |  |
| AIC | 3299071 |  | 3298758 |  |  |  | 3427278 |  | 3426768 |  |  |  |
| Deviance (-2xlog-likelihood) | 15800250 |  | 15800987 |  |  |  | 21843901 |  | 21849139 |  |  |  |

Level 3 = intersectional strata (n=398); Level 2 = individuals (n=83,304); Level 1 = repeated measurement observations (n=483,887).
\* p<0.05 \*\* p<0.01 \*\*\*
p<0.001

In Model 2P, after adding the main effects and their interactions with age, the between-strata share of variability falls sharply. The intercept VPC drops from 18.45% to 3.76%, showing that the main effects explain most of the between-strata differences in centred-age health, with an intercept PCV of 82.73%. For age-related change, the slope VPC decreases from 15.37% to 9.15%, indicating that much of the between-strata heterogeneity in age slopes was explained by the age-by-main-effect interactions; the slope PCV of 44.48% shows that Model 2 reduces the stratum-level age-slope variance by almost half relative to Model 1.

Estimates at the centred age reveal patterned inequalities, with poorer physical health reported by women, Bangladeshi and Pakistani groups, and those in routine/manual or no-work categories. However, significant interaction terms indicate these gradients vary across age, with steeper physical health decline among women and those in routine or no work history categories.

Model 1M shows that mental health changes non-linearly with age, with both squared and cubic terms statistically significant, indicating a non-linear trajectory. Most of the variation is within individuals over time (52.08), followed by variation between individuals (46.23), with a smaller but still meaningful share attributable to intersectional strata (22.28). Heterogeneity in age-related change is minimal, with small age–slope variances of 0.009 at level 3 and 0.019 at level 2, suggesting that inequalities in mental health are driven more by differences in centred-age health than by differences in rates of decline.

In Model 2M, after adding main effects and their interactions with age, between-strata variation reduces substantially. The intercept VPC falls from 30.95% to 17.44%, indicating that a considerable proportion of between-strata centred-age differences is explained by the additive effects. The intercept PCV of 84.32% suggests that most strata-level centred-age variation is accounted for. For age-related change, the slope VPC drops from 32.52% to 7.01%, and the slope PCV of 53.62% indicates that Model 2 reduces the stratum-level age-slope variance by over half relative to Model 1, after adding the stratum-defining main effects and their interactions with age. Together, these reductions show that the main effects and their age interactions capture a substantial share of intersectional inequalities in both centred-age levels and trajectories in mental health.

Estimates at the centred age reveal systematic inequalities by ethnicity, gender and occupational class: African and Indian groups have higher predicted mental health scores, while women and those with no work history have lower predicted scores. Because not all generations are observed at the centred age, generational differences are interpreted through the predicted trajectories in Figures 2 and 3, which are plotted only across observed age ranges (Supplementary Table S1). For physical health (SF-12 PCS), predicted scores decline steadily with age and confidence intervals largely overlap where adjacent generations coincide, suggesting stronger age than generational patterning. For mental health (SF-12 MCS), later-born generations show lower predicted scores within their observed age ranges. However, because age overlap is limited mainly to adjacent generations, these patterns are interpreted as descriptive model-based contrasts. The Boomer generation shows a relatively flat trajectory across its observed age range, while Generation Z has lower predicted mental health in early adulthood.

**Figure 2.**
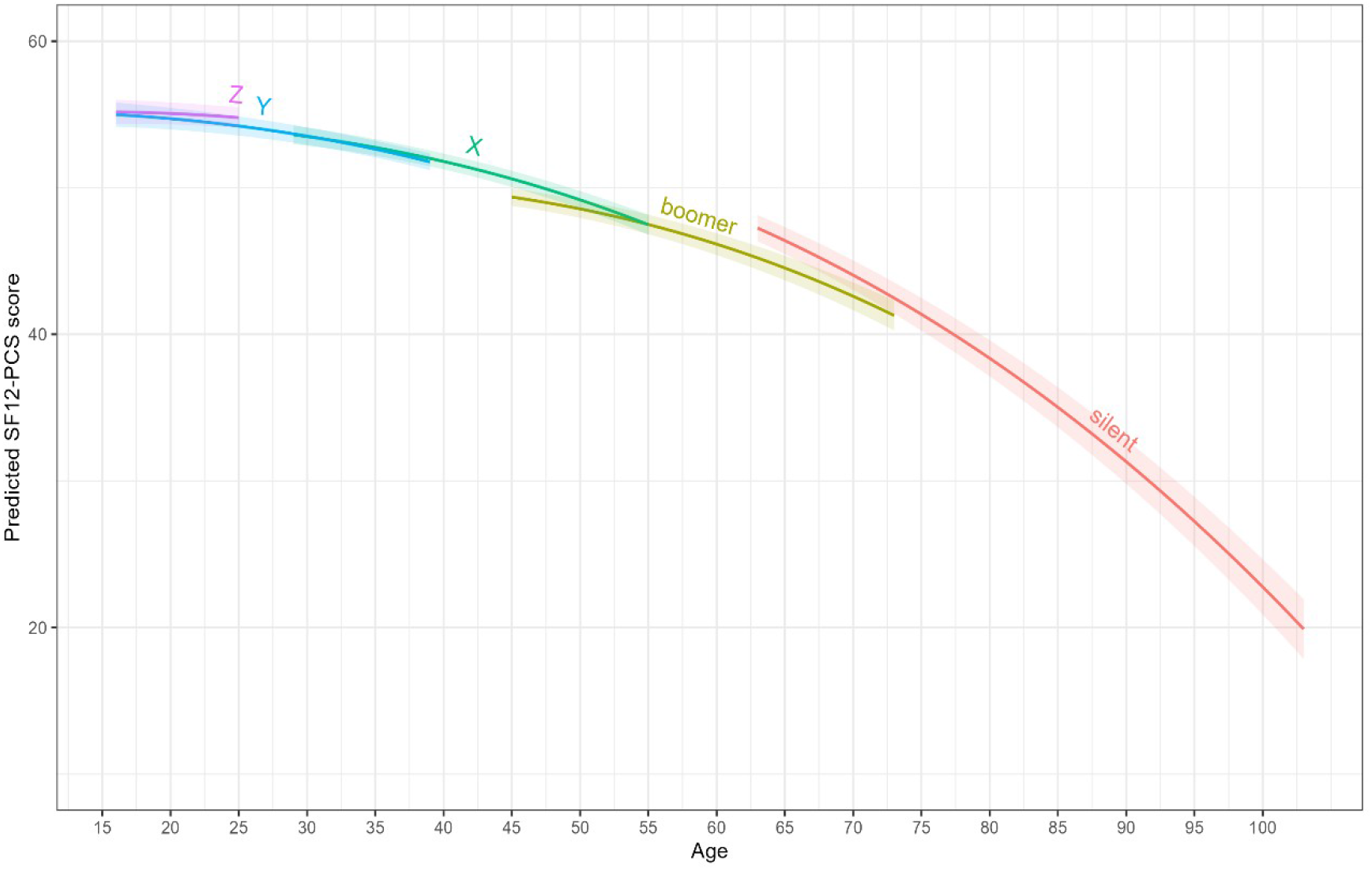
Generational Effects SF-12 PCS score.

**Figure 3.**
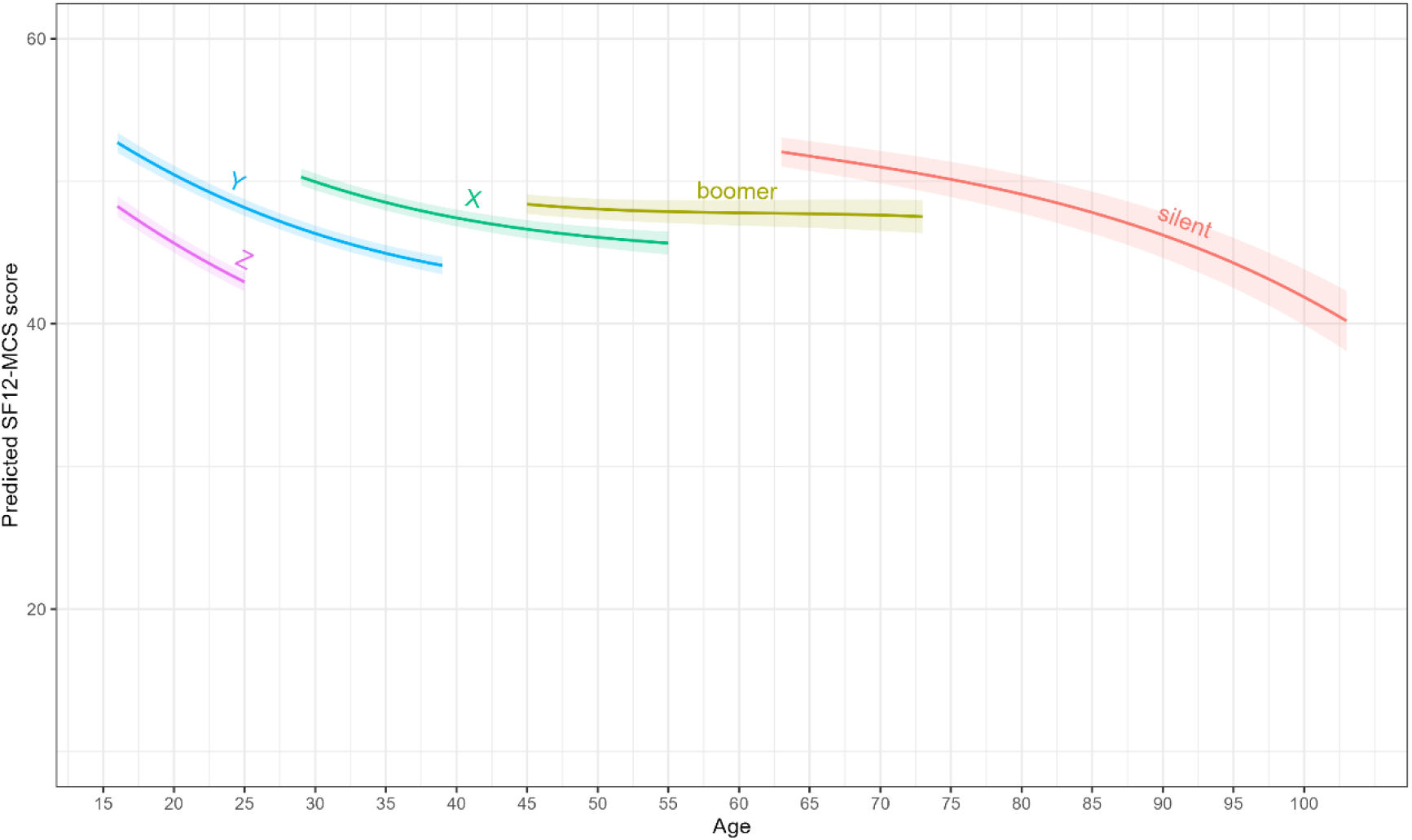
Generational Effects SF-12 MCS.

Variance function plots (Supplementary Figures S1 and S2) show that for physical health, between-individual variance increases sharply with age, consistent with cumulative disadvantage widening inequalities over the life course, whereas for mental health the pattern is flatter and overall variance lower. In both cases, between-strata variance is smaller than between-individual variance and reduced further in Model 2, indicating that most heterogeneity remains within rather than between intersectional strata.

Figure 4 suggests that intersectional inequalities in physical health amongst the most advantaged groups are wider in older generations, with favourable trajectories appearing more compressed among later-born cohorts. White British and Other White managerial and professional men tend to sit towards the upper end of the physical-health distribution relative to the other strata. In Generation Z, White Irish managerial and professional women have among the most favourable predicted physical health trajectories. For the Silent generation, White Irish, Other White and Mixed men appear higher in the fitted lines, though these differences are uncertain and may not be statistically meaningful given likely confidence-interval overlap. In terms of gender differences, White British managerial and professional men generally have higher predicted physical health than women, with this gap appearing wider amongst older generations.

**Figure 4.**
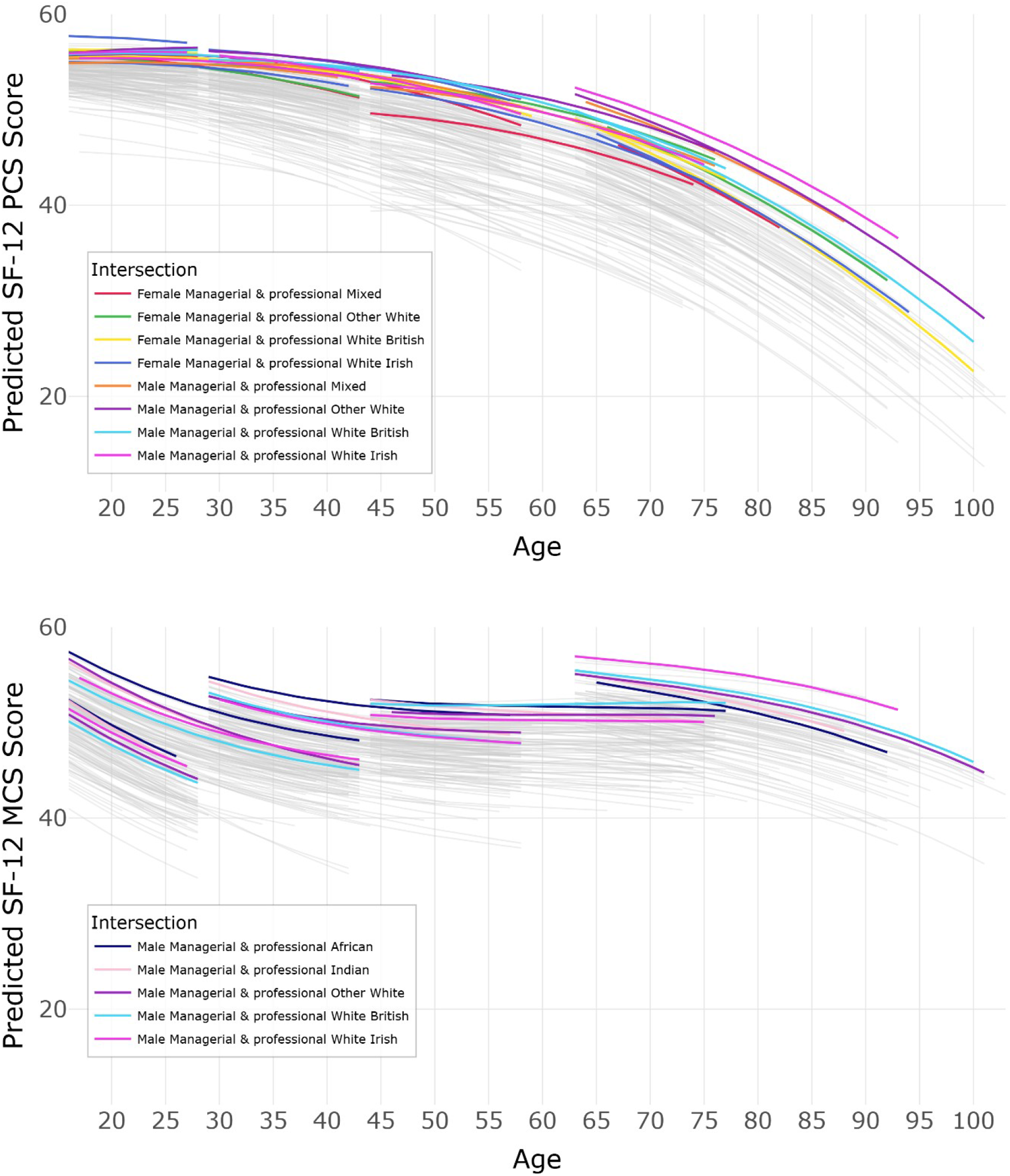
Advantaged Intersectional Trajectories in Physical and Mental Health in the UK (2009-2024)

For mental health, the picture is less consistent than for physical health: White British managerial and professional men exhibit a clearer relative advantage only among the Boomer generation, while in other generations the advantage shifts to different intersections. Across generations, several managerial and professional male intersections – particularly White Irish, Other White, African and Indian – sit towards the upper end of the predicted mental-health distribution. Differences between these highlighted groups are generally modest, and their trajectories often appear similar in shape and level, indicating a grouping of comparatively favourable outcomes. Notably, in Generations X and Y, Black African and Indian male trajectories often sit above the White British reference, effectively reversing the ethnic hierarchy seen in physical health. Some generational variation is evident, with White Irish men advantaged amongst the Silent generation, White British men among the Boomer generation, and African, Indian and Other White men tending to sit towards the upper range in later-born cohorts.

Overall, these patterns suggest that, within the modelled age ranges, advantage among managerial and professional groups is more differentiated in older generations, while in younger generations favourable outcomes are more compressed and shift across intersections rather than forming a stable hierarchy.

Figure 5 examines intersections occupying the least healthy end of the distribution, focusing on individuals with no work history. For physical health, the disadvantaged intersections vary across generational age ranges. In the Silent and Boomer generations, Bangladeshi and Pakistani women with no work history tend to sit towards the lower end of the predicted physical-health distribution. In Generation Z, trajectories are more tightly clustered overall, suggesting less differentiation among young people outside the labour market, although Bangladeshi women and Pakistani men continue to sit towards the lower end of the distribution. In Generation Y, Bangladeshi men and White Irish women appear among the more disadvantaged intersections, while in Generation X, White British men and women sit nearer the lower end of the distribution, with the latter showing a particularly steep decline with age.

**Figure 5.**
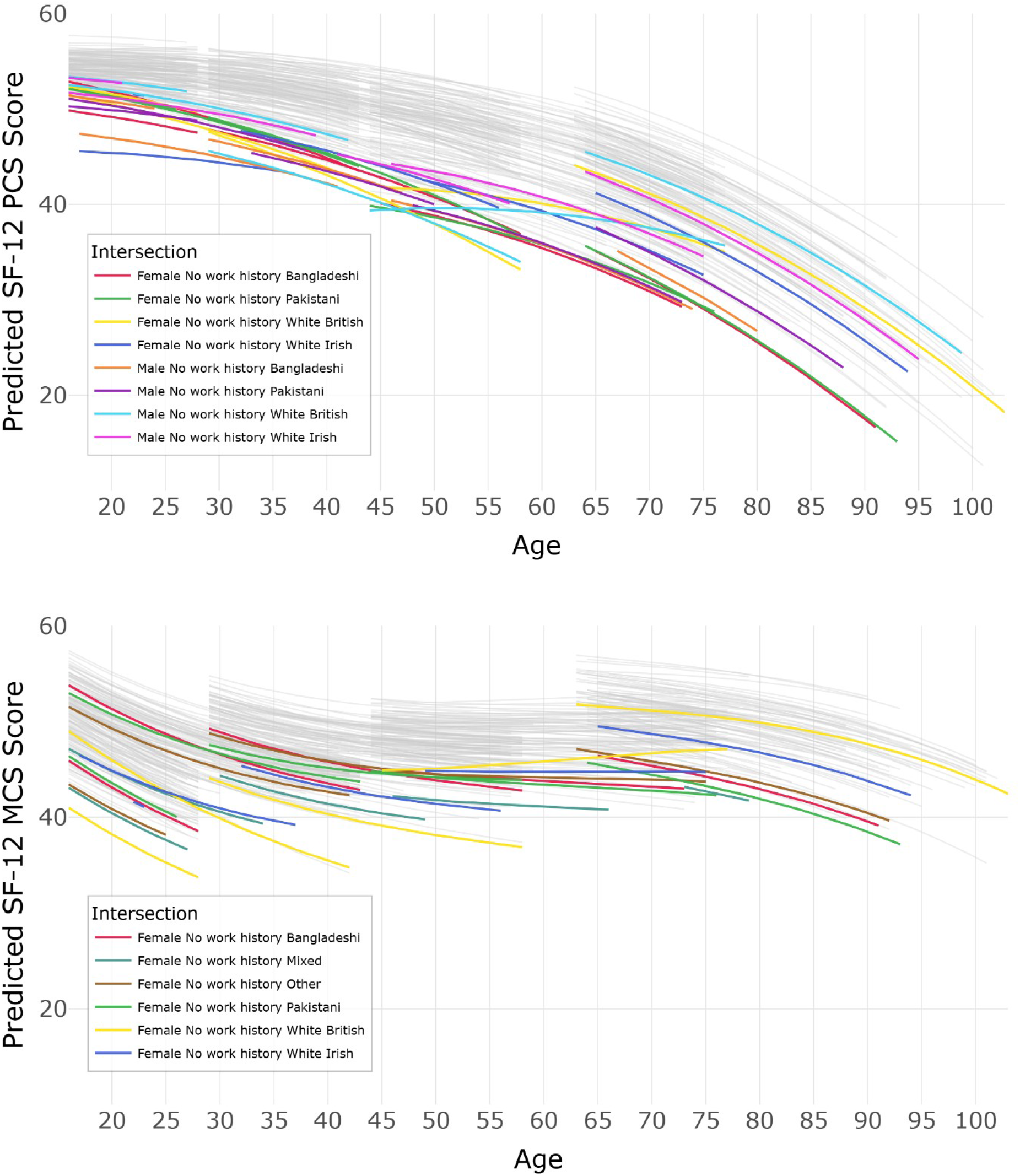
Disadvantaged Intersectional Trajectories in Physical and Mental Health in the UK (2009-2024)

For mental health, patterns are less stable. Women with no work history generally sit towards the lower end of the predicted mental-health distribution across generations, and male intersections are therefore not shown. In the Silent and Boomer generations, Pakistani, Bangladeshi and Other-ethnicity women tend to appear among the more disadvantaged groups. In Generations X, Y and especially Z, White British women display comparatively poor trajectories, with a noticeably steep decline in Generation Y and a particularly pronounced disadvantage in Generation Z. This trajectory falls below several other ethnic groups within the observed age range, contrasting with the relative advantage seen for White British groups in some older cohorts. Women of Mixed and Other ethnicity also occupy the lower end of the distribution in some generations. Overall, these intersections form a cluster of disadvantaged trajectories, with shifts in which groups appear most affected across generations rather than a stable hierarchy.

Taken together, these patterns across physical and mental health suggest that weak labour-market attachment is consistently associated with disadvantage, while the intersections most affected vary across generations rather than following a fixed hierarchy.

## Discussion

Our novel study found complex, intersectional inequalities in trajectories in both physical and mental health, underlining that health outcomes are jointly shaped by ethnicity, gender, social class, and generation in the UK. Physical health inequalities widened with age, consistent with cumulative disadvantage, whereas poorer mental health was more evident among later-born cohorts observed at younger ages during the study period. These findings align with the concept of unequal ageing, which emphasises the ways in which ageing processes are structured by intersecting inequalities (Holman & Walker, 2020). In line with intersectionality theory (Collins, 2002; Crenshaw, 1989, 1991) and extended in health research by Bauer (2014) and Bowleg (2012), no single axis of inequality fully explained the observed health inequalities.

Advantages and disadvantages in health are distributed unevenly in unexpected ways. For instance, White British men in managerial and professional occupations – a conventionally privileged intersection – had better self-rated physical health trajectories across most generations, but not uniformly better mental health. This advantage was clearest amongst the Boomer generation; in other generational age ranges, some ethnically minoritised groups had more favourable predicted mental health trajectories. Importantly, intersectional patterns in physical health were not mirrored in mental health. Intersectional differences did not simply amplify generational differences; instead, they redistributed them across gender, ethnicity and occupational-class positions. This divergence suggests that different mechanisms and pathways shape these outcomes, and that advantages or disadvantages in one domain cannot be assumed to generalise to the other.

### Generational patterning in physical and mental health trajectories

Overall generational differences in physical health were modest: within overlapping portions of the age range, physical-health differences between generations were small and confined to particular age ranges. This aligns with evidence from Europe and the United States showing poorer physical health among Baby Boomers – attributed to factors including ‘changing associations between disability and chronic disease morbidity, multimorbidity, and obesity’ (Gimeno et al., 2024:8).

Clearer generational patterning was observed for mental health, where later-born cohorts had lower predicted SF-12 MCS scores in the modelled trajectories. This pattern is consistent with previous research finding Baby Boomers’ mental health improved over time (Bell, 2014; Bell et al., 2024). Debate remains over whether poorer mental health among later-born cohorts reflects greater exposure to structural and psychosocial risks, increased reporting, or both (Foulkes & Andrews, 2023; McGorry et al., 2025). In our analysis, lower SF-12 MCS scores were not evenly distributed across later-born cohorts, but varied by gender, ethnicity, and occupational class. Because SF-12 comparability across generations is assumed rather than directly tested, these patterns should be interpreted cautiously.

### Intersectional inequalities in trajectories

Intersectional advantage and disadvantage varied across generations rather than forming a stable hierarchy. The fact that White British managerial and professional men do not consistently sit at the upper end of the physical-health distribution in Generation Z and the Silent Generation suggests that the compounded advantage of class and gender is unstable across generations. This instability was especially apparent for mental health, where African, Indian, Other White and White Irish men often showed more favourable trajectories than the White British majority. While this could reflect a ‘healthy immigrant’ effect (Dorsett et al., 2019), such advantages typically attenuate with duration of residence due to acculturation and the stressors of a hostile environment. Given this potential decay, and the fact that the UK’s foreign-born share remains modest (∼16 %), these favourable outcomes likely also reflect distinct protective mechanisms such as community cohesion (Fernando, 2010) or cultural identity. Further, mental health processes are more socially contingent than physical health, mediated by factors such as identity and stress (Thoits, 2013), stigma (Bharadwaj et al., 2017), and social comparison (Arigo et al., 2024). This is highly relevant to an intersectional perspective, as it underlines how lived experience associated with social location – shaped by recognition, discrimination, and exclusion – can produce shifting patterns of mental health advantage and disadvantage across generations (Burgess et al., 2025).

Most striking was the poorer predicted mental health of White British groups in later-born cohorts. White British women with no work history tended to sit towards the very lowest end of the mental-health distribution from Generation X onwards, with poorer mental health in Generation Y and especially Generation Z. This is striking given the relative advantage observed for some White British groups in older cohorts. One possible interpretation, drawing on life course research linking mothers’ employment-marriage biographies to mid-life health, is that no work history may capture a set of family, caregiving, partnership, and labour-market circumstances associated with poorer mental health (McDonough et al., 2015). A second possibility is that these patterns reflect changing socioeconomic expectations and insecurity among groups whose expected position has become less secure (Case & Deaton, 2020). More broadly, White British women in Generation Z show comparatively poor predicted mental health across occupational class groups, although whether this reflects differences in reporting, psychosocial risk, labour-market insecurity, family/care circumstances, or other unmeasured mechanisms cannot be determined here. This suggests a need to consider whether existing mental health support reaches groups with poorer predicted mental health, while avoiding overly narrow targeting based on any single category. In addition, White British women with no work history have among the worst physical health trajectories in younger generations. With respect to gender differences, men at each level of occupational class generally had a slight predicted mental health advantage over women in all cohorts, with the gap appearing more pronounced in Generation Z. This pattern is consistent with evidence that social media use is more strongly linked to worsening well-being among adolescent girls than boys, with higher levels of use predicting declines in happiness and greater socio-emotional difficulties over time (Booker et al., 2018). However, this male ‘advantage’ should be interpreted with caution. It reflects the well-established ‘gender paradox’ in mental health, where men report lower levels of internalising symptoms despite exhibiting higher rates of suicide and externalising disorders (Canetto & Sakinofsky, 1998).

Our findings revealed significant persistent disadvantage among Bangladeshi and Pakistani groups, especially those with no work history. Both men and women in these groups sat at the lower end of the physical-health distribution, showing pronounced cumulative disadvantage. This aligns with disparities in limiting long-term illness (Bécares, 2015) and co-occurring diseases in children (Hesketh, 2016). However, this morbidity disadvantage contrasts with ONS estimates showing high life expectancy for Bangladeshi groups (Office for National Statistics, 2021). This paradox may reflect differences in data sources, sampling, time periods, and the influence of selective migration and reporting effects. This mirrors the ‘morbidity-mortality paradox’ often seen in gender research, suggesting that while some groups live longer, they endure a greater burden of ill health, potentially exacerbated by structural barriers in healthcare access and experience (Kapadia et al., 2022). Crucially, ethnicity is not a biological risk factor, but a proxy for complex migration histories; while the ‘healthy migrant’ effect may confer a mortality advantage, it does not necessarily protect against the accumulation of chronic morbidity and disability in the UK labour market. Notably, the intersections most disadvantaged for physical health were not always the same as those disadvantaged for mental health. Especially in younger generations, White British and Mixed groups (particularly women with no work history) had worse mental health trajectories than their Bangladeshi and Pakistani counterparts. This diverges from earlier UK research suggesting elevated psychological distress among these groups (Dorsett et al., 2019). Finally, Indians had better physical health trajectories than Bangladeshis and Pakistanis. This cautions against the common practice of statistically grouping South Asians together (Hayanga et al., 2023).

### Structural Inequalities and Health Trajectories

The observed trajectories are consistent with health inequalities unfolding within structural systems of power. In the UK, ethnically minoritised populations have long faced inequities in experience of health services driven by discrimination, and structural and environmental disadvantage (Robertson et al., 2021). Persistent disadvantage among Pakistani and Bangladeshi groups likely reflects the enduring effects of institutional racism, labour market exclusion and exploitation, and neighbourhood deprivation (Bécares, 2015; Nazroo, 2003; D. R. Williams et al., 2019). At the same time, the emergence of poor mental health among young White British women highlights how inequality and discrimination is not always worse for ethnically minoritised groups across all outcomes. As with structural racism, structural sexism can shape health inequalities through institutional norms that restrict economic participation, caregiving expectations, and access to care. Homan et al. (2021) demonstrated that sexism and racism operate jointly to shape health risks, with the most adverse outcomes occurring for women at the intersection of multiple forms of structural oppression. Alongside racism and sexism, our analysis also clearly emphasises the potential role of ‘classism’ - occupational class fixed effects were more pronounced than those for gender or ethnicity, strongly suggesting that an intersectionality discourse focussed only on racism and sexism misses a critical foundation and fundamental cause of health inequalities (Link & Phelan, 1995). At the same time, class differences are variegated by other axes of inequality, cautioning against a simplistic view of class inequality.

Our findings of shifting mental health disadvantage across generational age ranges, alongside widening physical health inequality with age, align with the idea that historical exposures and changing social environments shape health trajectories. Life course epidemiology highlights that early-life socioeconomic disadvantage has a ‘long reach’, influencing adult health through cumulative biological wear and social disadvantage (Kuh et al., 2025). The patterns we observed may reflect changes in labour markets, educational regimes, digital environments, and welfare systems, alongside the shifting dynamics of structural systems of discrimination. Existing research suggests that equity-oriented health policies must tackle systematic inequalities in exposure to different social, commercial and policy determinants of health over time (Bambra et al., 2021; Gilmore et al., 2023; Marmot et al., 2020), whilst also in the shorter term directly addressing the needs of the intersections experiencing the poorest outcomes. Future work should examine how different policies or institutional practices impact different intersectional groups over time to inform proportionate universalist approaches that combine universal provision with targeted intensification according to intersecting forms of disadvantage (Carey et al., 2015).

## Limitations

Generational comparisons are limited by the 2009–2024 observation window, which captures generations at different ages and contexts. Comparisons between non-adjacent generations should therefore be interpreted as descriptive model-based contrasts rather than fully age-matched cohort comparisons. Our results cannot separate out age, cohort and period effects. Although the analysis engages with life course theory by examining age-based trajectories and patterns consistent with cumulative disadvantage, it does not directly model early-life exposures, critical periods, chains of risk, or the mechanisms through which disadvantage accumulates. Future research could build on this descriptive foundation to examine these mechanisms more directly. We did not incorporate UKHLS survey weights, stratification, clustering, or household-level clustering. This reflects current limitations of longitudinal MAIHDA and the computational demands of the random-slope model. As a result, the results may not be fully representative of the UK population. Key demographic and socioeconomic variables related to differential selection and non-response are included as stratum-defining factors, but this is not equivalent to design-weighted estimation. NS-SEC was also treated as a person-level stratum-defining characteristic, although occupational class can change over time and may itself be affected by health. Future work could use cross-classified or dynamic specifications to examine changing socioeconomic position more directly (Bell et al., 2024). The granularity of our analysis is both a strength and limitation: while we highlight intersectional differences across physical and mental health trajectories, space restrictions necessarily limit more nuanced discussion. A further limitation is that intersectional differences may partly reflect measurement bias. For example, Galenkamp et al. (2018) found that while SF-12 was measurement invariant for gender, age and education, adjustment for differential item functioning reduced ethnic inequalities in physical health, and to a lesser extent, mental health. Therefore, our findings on ethnicity must be interpreted with caution. We also assume that SF-12 PCS and MCS are sufficiently comparable across generations; cohort differences in symptom recognition, expectations of health, or willingness to report distress may affect comparability, particularly for mental health. Additionally, while systems of discrimination undoubtedly contribute to health inequalities, a range of factors are implicated - including genetic, biological, behavioural, migration history and spatial - not all of which constitute discrimination; without analysing these determinants, we are limited in the claims we can make. Finally, ∼13% of observations were excluded due to non-completion or missing data, and non-random attrition may introduce selection or survivor bias. Excluding individuals observed only in education or training may also disproportionately affect later-born cohorts.

## Conclusion

Our analysis provides novel longitudinal insights into age-related health inequalities, showing that single-axis approaches to inequality obscure population heterogeneity. It highlights several unanticipated intersections with particularly stronger or weaker physical and mental health trajectories, underscoring the value of examining health inequalities through an intersectional lens. Methodologically, the novel use of longitudinal MAIHDA models demonstrates the value of modelling trajectories to capture both between- and within-group heterogeneity in health over time. The findings show that age-related health trajectories are not uniform, but vary across intersecting social positions and historically situated generations. Taken together, the results emphasise the value of combining intersectional and life course perspectives for understanding and addressing health inequalities. The results caution against simple targeted intervention based on single categories. Instead, the findings are consistent with a framework of ‘proportionate universalism’ (Carey et al., 2015), combining universal support with greater attention to groups whose predicted trajectories sit lower, or decline more steeply, across the observed age range, such as young women with no work history and older ethnically minoritised groups experiencing cumulative physical-health disadvantage. Future research should build on these findings to examine the mechanisms through which intersecting inequalities shape ageing trajectories and inform equitable policy interventions.

## Ethics statement

The Department of Sociological Studies ethics review panel, University of Sheffield, gave ethical approval for this work (application reference 051615) as part of the project “Storying Life Courses for Intersectional Inclusion: Ethnicity and Wellbeing across time and place”. The study is a secondary analysis of anonymised data from the UK Household Longitudinal Study, accessed via the UK Data Service under End User Licence; all data collection and participant consent procedures were managed by the original study team.

## Data availability

This study uses data from the UK Household Longitudinal Study (Understanding Society), available to registered researchers from the UK Data Service (SN 6614) under End User Licence. Technical documentation is available at https://github.com/danjholman/IntersectExplorer-Shiny and the interactive application at https://inequalities-research.shinyapps.io/IntersectExplorer/.

## Author contributions

Daniel Holman: Conceptualisation, Methodology, Formal analysis, Visualisation, Funding acquisition, Writing – Original Draft. Andrew Bell: Conceptualisation, Methodology, Formal analysis, Funding acquisition, Writing – Reviewing and Editing. Matthew Bennett: Conceptualisation, Funding acquisition, Writing – Reviewing and Editing. Helena Mendes Constante: Conceptualisation, Funding acquisition, Writing – Reviewing and Editing. Mark Green: Conceptualisation, Funding acquisition, Writing – Reviewing and Editing. Aneta Piekut: Conceptualisation, Funding acquisition, Writing – Reviewing and Editing.

## Acknowledgements

This paper has been presented to audiences at the following fora, whom we wish to thank for their constructive and insightful feedback: Lake Como School of Advanced Studies (’Transitions in Ageing’ workshop, 2025); Bocconi University, Italy (2025); the Society for Epidemiologic Research 2025 Annual Conference in Boston; and a Centre for Ageing Better seminar (2025).

## Declaration of generative AI and AI-assisted technologies in the manuscript preparation process

During the preparation of this work, the authors used ChatGPT for assistance with developing the accompanying interactive application and for improving the clarity and readability of the manuscript text. The authors reviewed and edited the output as needed and take full responsibility for the content of the published article.

## Modelling approach – Technical details

A key feature of the longitudinal MAIHDA approach is that the variance at each level is allowed to vary with age. The total stratum-level variance captures the extent of intersectional inequalities, and shows how these widen or narrow across the life course. This can be meaningfully compared to the variance observed within strata but between individuals, and to the variance within individuals across occasions (levels 2 and 1, respectively). In this way, the extent to which strata are predictive of health trajectories relative to other sources of individual- and occasion-level variability is assessed. Due to the variance functions in longitudinal MAIHDA changing with age, it is not possible to calculate a single overall Variance Partitioning Coefficient (VPC), as in cross-sectional MAIHDA. Instead, two separate VPCs are calculated. The intercept VPC quantifies the proportion of variance in health at the mean age (the intercept) that is explained by intersectional strata. It was calculated by dividing the stratum-level intercept variance by the total intercept variance (i.e. stratum-level plus individual-level). The slope VPC quantifies the proportion of variance in rates of change with age (the slopes) that is explained by intersectional strata. It was calculated by dividing the stratum-level slope variance by the total slope variance (i.e. stratum-level plus individual-level). Together, these VPCs show how much of the between-individual heterogeneity in starting points and trajectories of health ageing can be attributed to intersectional inequalities.

Model 2 expands Model 1 by adding the main effects of the intersection-defining variables, in this case gender, ethnicity, NS-SEC, and generation, and their interactions with age. This enables estimation of both differences in health at the mean age (intercepts) and differences in age-related trajectories (slopes) across strata. The random part of the model is the same as Model 1, but because additive fixed effects and their interactions are included, some of the stratum-level variance is expected to be explained by them. Model 2 therefore also assesses the extent to which variability in the age slope can be explained by these additive interactions with age – that is, additive effects on the size of the age effect – which are operationalised through two-way interactions between age and the intersection-defining variables.

Variance functions were calculated at the stratum level to show how much variance remained after accounting for additive intersectional effects, and how this varied with age. Comparing variance functions for Model 1 and Model 2 indicates whether intersectional inequalities are largely additive (if strata-level variance drops substantially) or whether substantial unexplained variance remains, suggesting multiplicative or unique intersectional effects. As with Model 1, we calculated intercept and slope VPCs for Model 2, quantifying the proportion of variance in mean-age health and trajectories that was explained by intersectional strata. Finally, we calculated the Proportional Change in Variance (PCV) of the strata-level intercepts and slopes between models 1 and 2. Intercept PCVs showed how much of the stratum-level differences in mean-age health are explained by the additive intersectional variables, while slope PCVs show how much of the differences in age trajectories are explained by additive (age-by-variable interaction) terms.

**Table S1.** Cross-classification of modal and first observed NS-SEC.

| Modal NS-SEC | First observed managerial and professional | First observed intermediate | First observed routine and manual | First observed no work history |
| --- | --- | --- | --- | --- |
| Managerial and professional | 22,466 (86.6%) | 1573 (6.1%) | 1913 (7.4%) | 0 (0%) |
| Intermediate | 1681 (9.5%) | 13,789 (78.0%) | 2221 (12.6%) | 0 (0%) |
| Routine and manual | 1527 (4.8%) | 1617 (5.1%) | 28,853 (90.2%) | 0 (0%) |
| No work history | 0 (0%) | 0 (0%) | 0 (0%) | 7,664 (100%) |

Percentages are row percentages. Overall, 87.4% of individuals were classified in the same NS-SEC category under the modal and first-observed specifications.

**Table S2.** Comparison of selected robustness model estimates.

| Estimate | Main PCS | First-observed PCS | Main MCS | First-observed MCS |
| --- | --- | --- | --- | --- |
| Managerial and professional | Reference | Reference | Reference | Reference |
| Intermediate | 1.498 | -1.346 | -.361 | -0.454 |
| Routine and manual | -3.475 | -3.408 | -1.162 | -1.069 |
| No work history | -7.868 | -7.849 | -4.514 | -4.496 |
| Age x Managerial and professional | Reference | Reference | Reference | Reference |
| Age x Intermediate | -0.001 | 0.000 | -.001 | 0.007 |
| Age x Routine and manual | -0.044 | -0.041 | 0.001 | 0.008 |
| Age x No work history | -0.096 | -0.094 | -0.042 | -0.033 |
| Strata intercept variance | 2.222 | 2.192 | 3.493 | 3.003 |
| Strata age-slope variance | 0.004 | 0.004 | 0.004 | 0.004 |

Estimates are selected fixed effects and variance components from Model 7. Managerial/professional is the reference NS-SEC category. Age interactions show differences in linear age slopes relative to the managerial/professional reference group. Full models included age, age<sup>2</sup>, age<sup>3</sup>, ethnicity, generation, NS-SEC, gender/sex, and interactions between age and each stratum-defining variable.

**Table S3.**
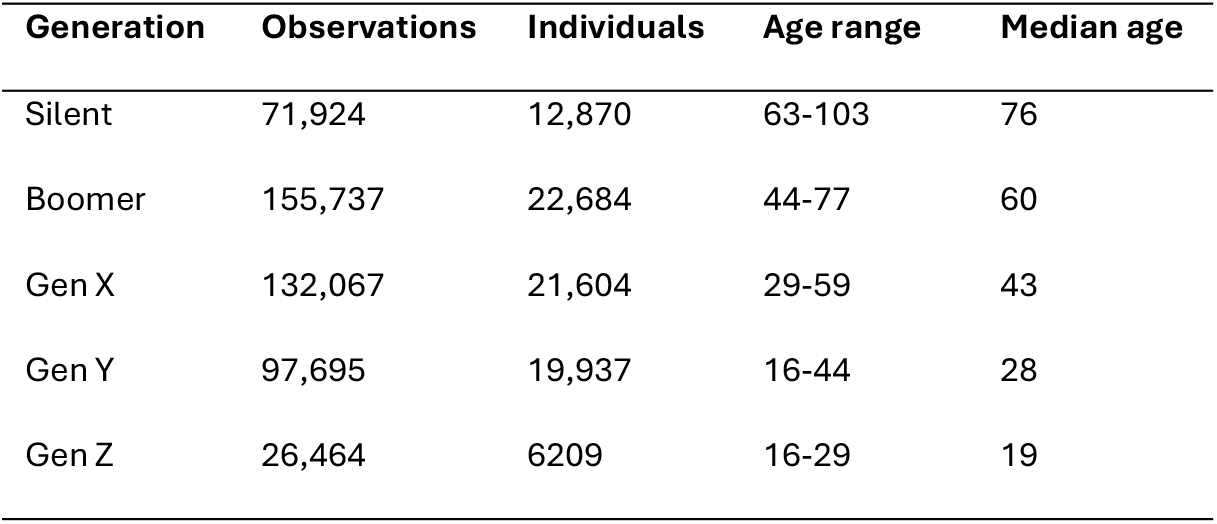
Age by generation.

| Generation | Observations | Individuals | Age range | Median age |
| --- | --- | --- | --- | --- |
| Silent | 71,924 | 12,870 | 63-103 | 76 |
| Boomer | 155,737 | 22,684 | 44-77 | 60 |
| Gen X | 132,067 | 21,604 | 29-59 | 43 |
| Gen Y | 97,695 | 19,937 | 16-44 | 28 |
| Gen Z | 26,464 | 6209 | 16-29 | 19 |

**Table S4.**
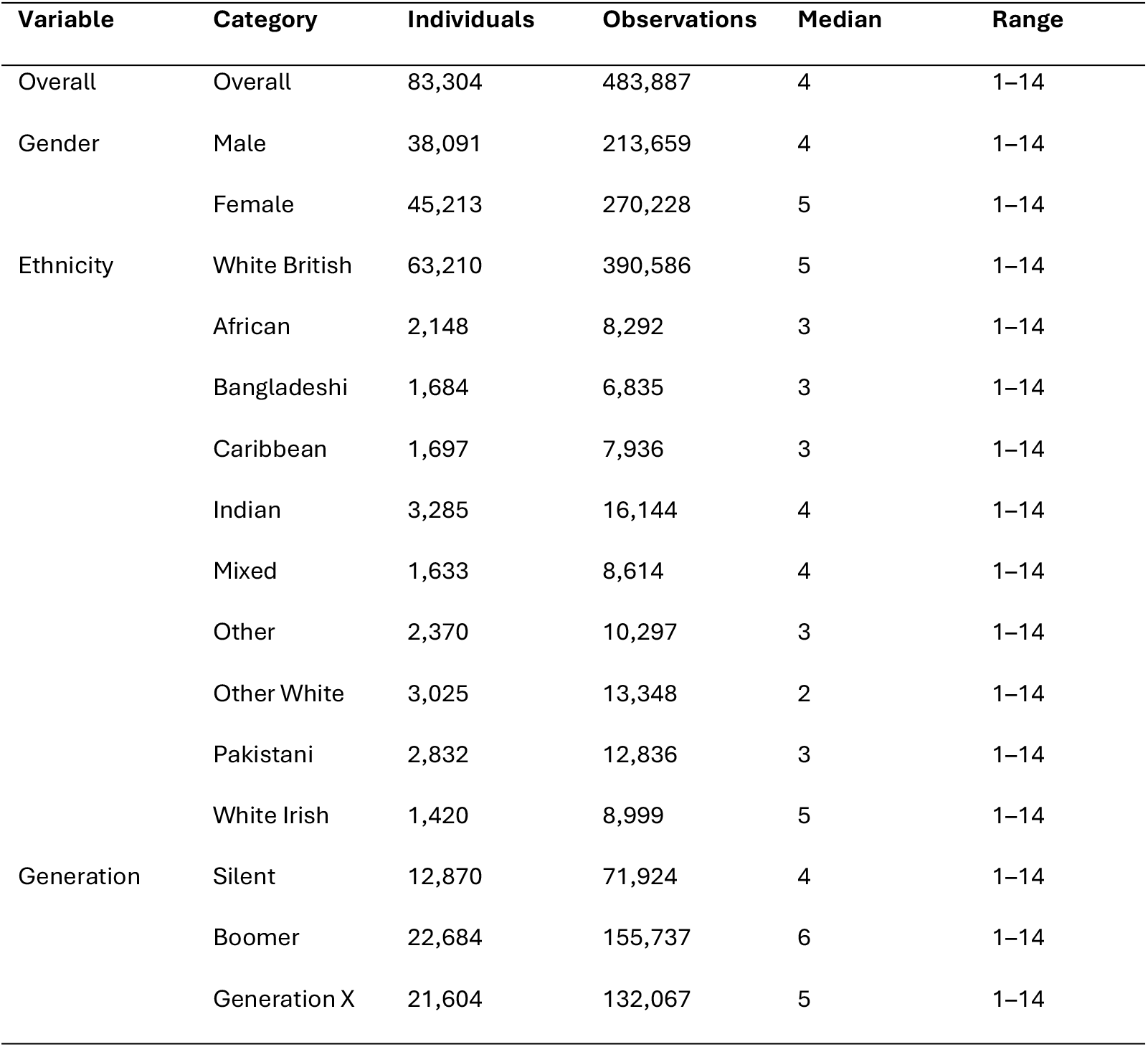
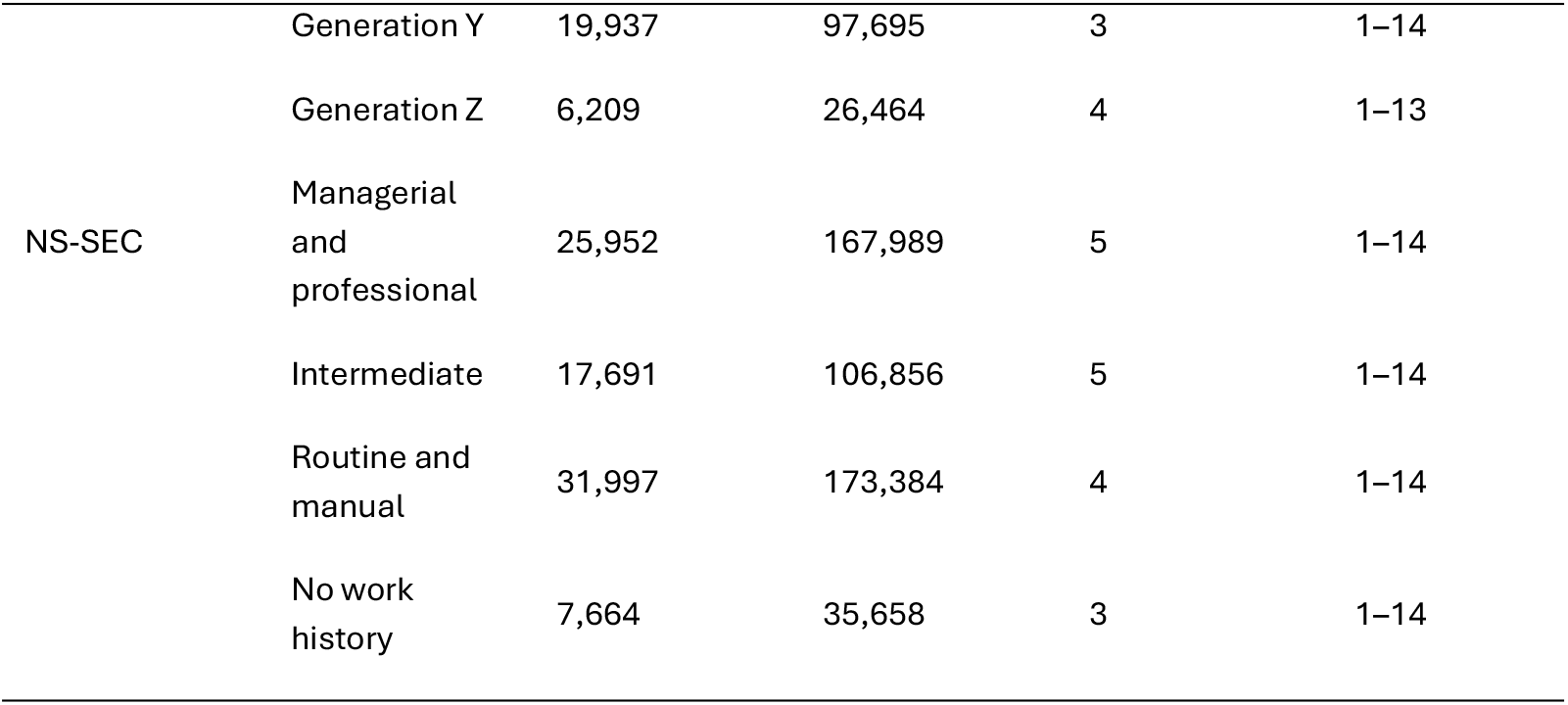
Repeated observations per individual in the analytical sample.

**Table S5.** All results, SF-12 PCS.

|  | Model A |  | Model B |  | Model C |  | Model D |  | Model 1 |  | Model E |  | Model 2 |  |  |  |
| --- | --- | --- | --- | --- | --- | --- | --- | --- | --- | --- | --- | --- | --- | --- | --- | --- |
|  | Estimate<br>s | SE | Estimate<br>s | SE | Estimate<br>s | SE | Estimate<br>s | SE | Estimate<br>s | SE | Estimate<br>s | SE | Estimate<br>s | SE | Estimate<br>s | SE |
| Intercept | 49.092**<br>* | .036 | 47.920**<br>* | .330 | 48.510**<br>* | .175 | 48.244**<br>* | .207 | 48.269**<br>* | .215 | 54.358**<br>* | .520 | 55.156**<br>* | .552 |  |  |
| Age (mean-centred) |  |  |  |  | -.241*** | .003 | -.273*** | .008 | -.274*** | .008 | -.273*** | .009 | -.215*** | .027 |  |  |
| Age squared |  |  |  |  | -.00385*** | .0000<br>7 | -.00387*** | .0001<br>6 | -.00437*** | .0001<br>6 | -.00440*** | .0001<br>6 | -.00513*** | .0002<br>6 |  |  |
| Age cubed |  |  |  |  | -.00002 | .0000<br>0 | -.00001 | .0000<br>0 | -.00003*** | .0000<br>0 | -.00003*** | .0000<br>0 | -.00002*** | .0000<br>1 |  |  |
|  |  |  |  |  |  |  |  |  |  |  |  |  | **Main effects** |  | **Age**<br>**interactions** |  |
| African (ref White British) |  |  |  |  |  |  |  |  |  |  | -.378 | .479 | -0.768 | .486 | -.034 | .022 |
| Bangladeshi |  |  |  |  |  |  |  |  |  |  | -<br>3.471*** | .506 | -<br>3.941*** | .524 | -.051* | .024 |
| Caribbean |  |  |  |  |  |  |  |  |  |  | -.764 | .487 | -1.035* | .484 | -.030 | .022 |
| Indian |  |  |  |  |  |  |  |  |  |  | -<br>1.576*** | .453 | -<br>1.495*** | .444 | .002 | .020 |
| Mixed |  |  |  |  |  |  |  |  |  |  | -.403 | .487 | -0.524 | .509 | -.005 | .023 |
| Other |  |  |  |  |  |  |  |  |  |  | -.932* | .467 | -1.221** | .463 | -.032 | .021 |
| Other White |  |  |  |  |  |  |  |  |  |  | .449 | .463 | 0.734 | .455 | .035 | .020 |
| Pakistani |  |  |  |  |  |  |  |  |  |  | -<br>3.165*** | .480 | -<br>3.465*** | .465 | -.040 | .021 |
| White Irish |  |  |  |  |  |  |  |  |  |  | -.183 | .487 | -0.138 | .484 | .009 | .021 |
| Baby Boomer (ref Silent Gen) |  |  |  |  |  |  |  |  |  |  | -<br>3.100*** | .517 | -<br>3.243*** | .519 | .085*** | .024 |
| Gen X |  |  |  |  |  |  |  |  |  |  | -<br>1.992*** | .499 | -<br>2.507*** | .526 | -.040 | .027 |
| Gen Y |  |  |  |  |  |  |  |  |  |  | -<br>1.833*** | .530 | -<br>3.027*** | .527 | -.070* | .032 |
| Gen Z |  |  |  |  |  |  |  |  |  |  | -.849 | .589 | -1.444 | .861 | -.027 | .040 |
| NS-SEC Intermed. (ref Manag. & pro.) |  |  |  |  |  |  |  |  |  |  | -<br>1.448*** | .313 | -<br>1.498*** | .319 | -.001 | .015 |
| Routine and manual |  |  |  |  |  |  |  |  |  |  | -<br>3.103*** | .313 | -<br>3.475*** | .303 | -.044** | .014 |
| No work history |  |  |  |  |  |  |  |  |  |  | -<br>7.011*** | .402 | -<br>7.868*** | .383 | -.096*** | .017 |
| Female (ref Male) |  |  |  |  |  |  |  |  |  |  | -<br>1.067*** | .236 | -<br>1.369*** | .233 | -.037*** | .011 |
| **Random effects** |  |  |  |  |  |  |  |  |  |  |  |  |  |  |  |  |
| Level 3 intercept variance |  |  | 39.680 |  | 8.952 |  | 11.820 |  | 12.862 |  | 2.821 |  | 2.222 |  |  |  |
| Level 3 age-slope variance |  |  |  |  |  |  | .008 |  | .008 |  | .007 |  | .004 |  |  |  |
| Level 3 intercept-slope covariance |  |  |  |  |  |  | .114 |  | .152 |  | .045 |  | .032 |  |  |  |
| Level 2 intercept var | 91.395 |  | 63.159 |  | 61.828 |  | 61.599 |  | 56.868 |  | 56.859 |  | 56.870 |  |  |  |
| Level 2 age-slope variance |  |  |  |  |  |  |  |  | .042 |  | .042 |  | .042 |  |  |  |
| Level 2 intercept-slope covariance |  |  |  |  |  |  |  |  | 1.201 |  | 1.200 |  | 1.201 |  |  |  |
| Level 1 variance | 40.257 |  | 40.230 |  | 38.873 |  | 38.798 |  | 37.945 |  | 37.944 |  | 37.945 |  |  |  |
| AIC | 3354141 |  | 3329361 |  | 3313197 |  | 3312506 |  | 3299071 |  | 3298750 |  | 3298758 |  |  |  |
| Deviance (-2xloglikelihood) | 1659720<br>4 |  | 1673194<br>9 |  | 1616230<br>5 |  | 1612781<br>9 |  | 1580025<br>0 |  | 1580033<br>6 |  | 1580098<br>7 |  |  |  |

Level 3 = intersectional strata (n=398); Level 2 = individuals (n=83,304); Level 1 = repeated measurement observations (n=483,887).
\* p<0.05 \*\* p<0.01 \*\*\*
p<0.001
This table presents the sequential model-building strategy for SF-12 PCS. Models A–E are intermediate models used to assess the age specification, random-effect structure, and addition of stratum-defining covariates. Models 1 and 2 are the main models reported in the manuscript.

**Table S6.**
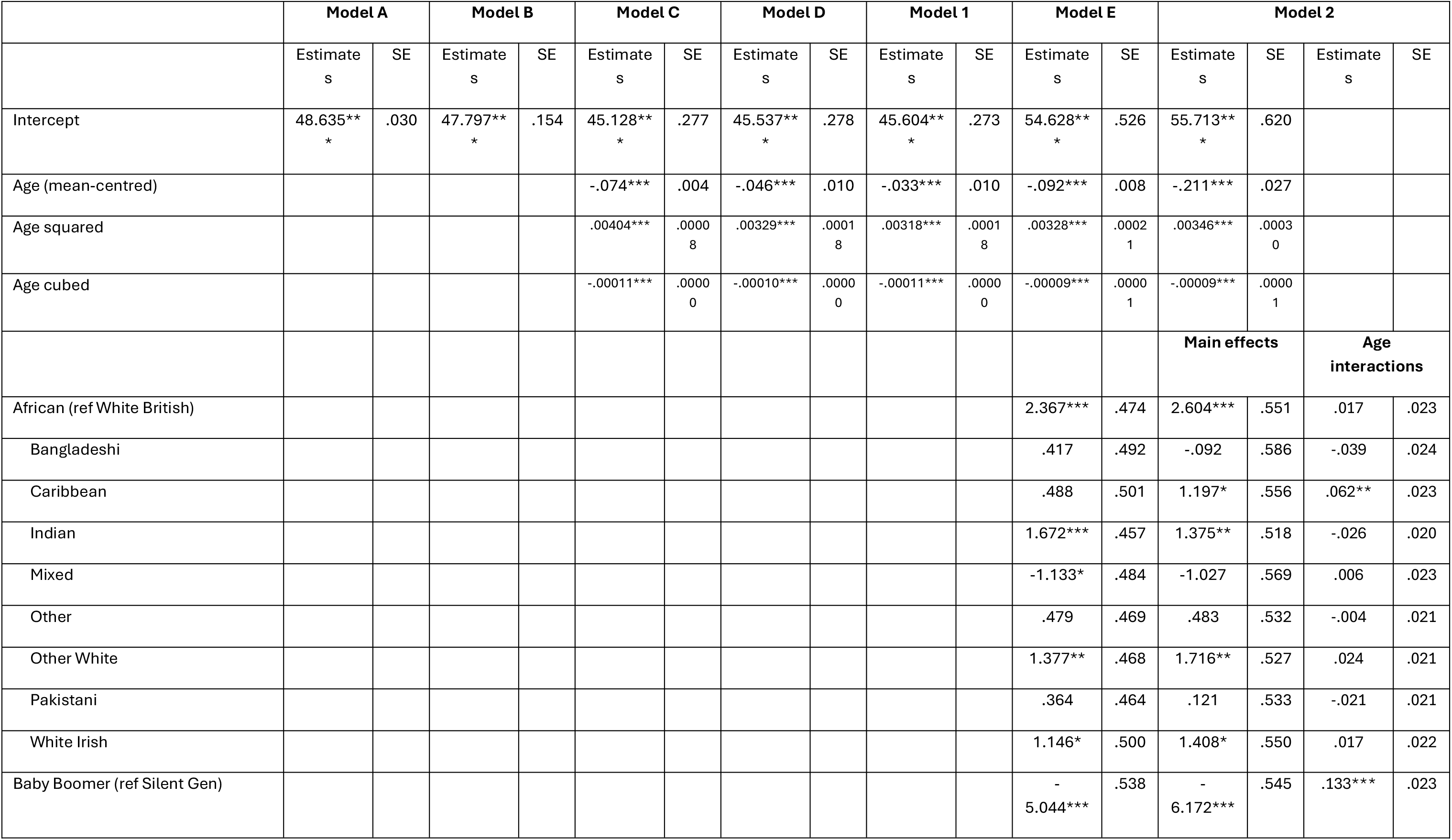
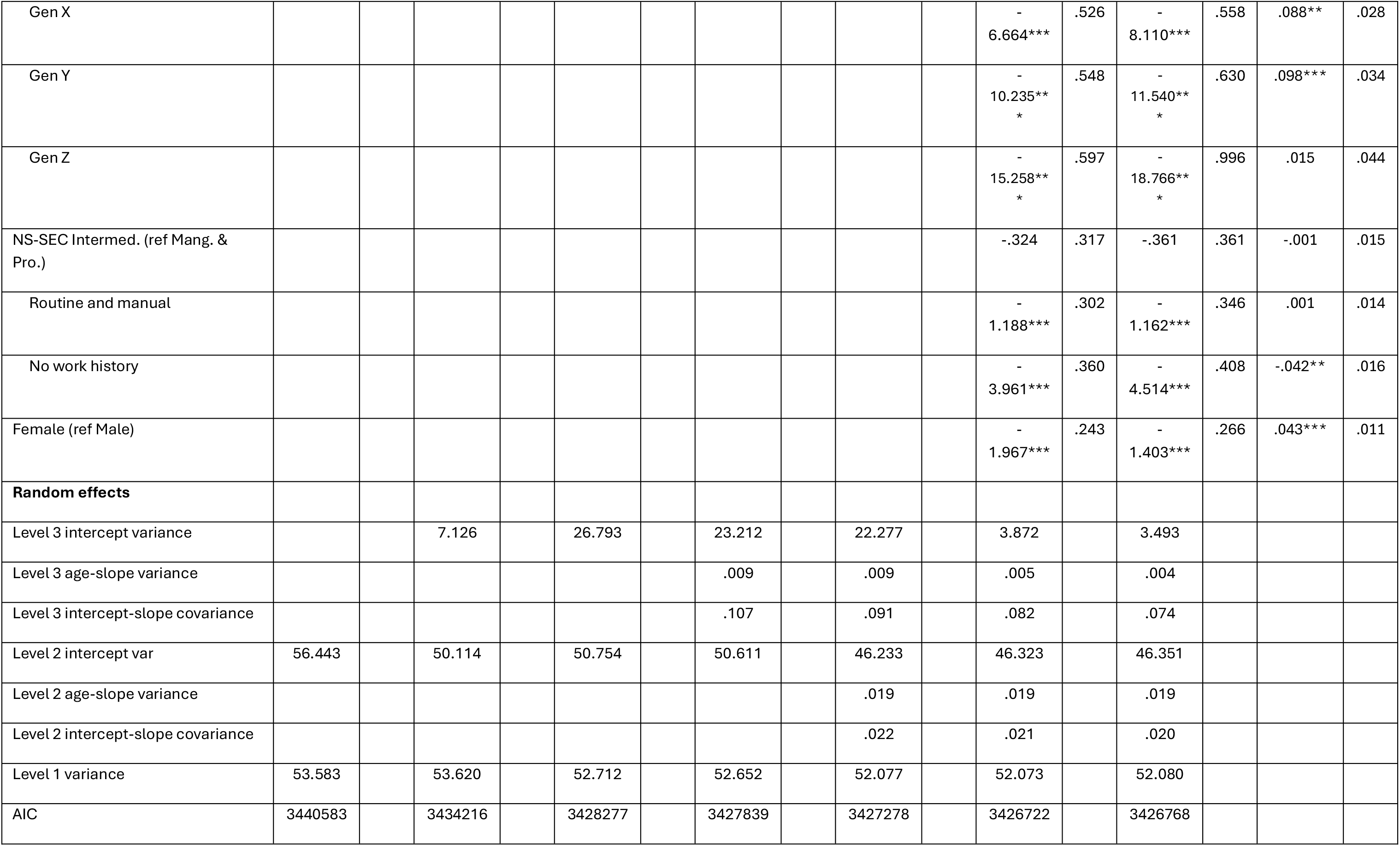
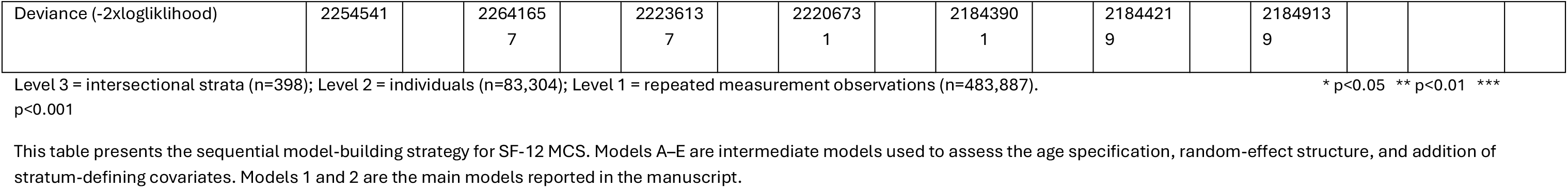
All results, SF-12 MCS.

**Figure S1.**
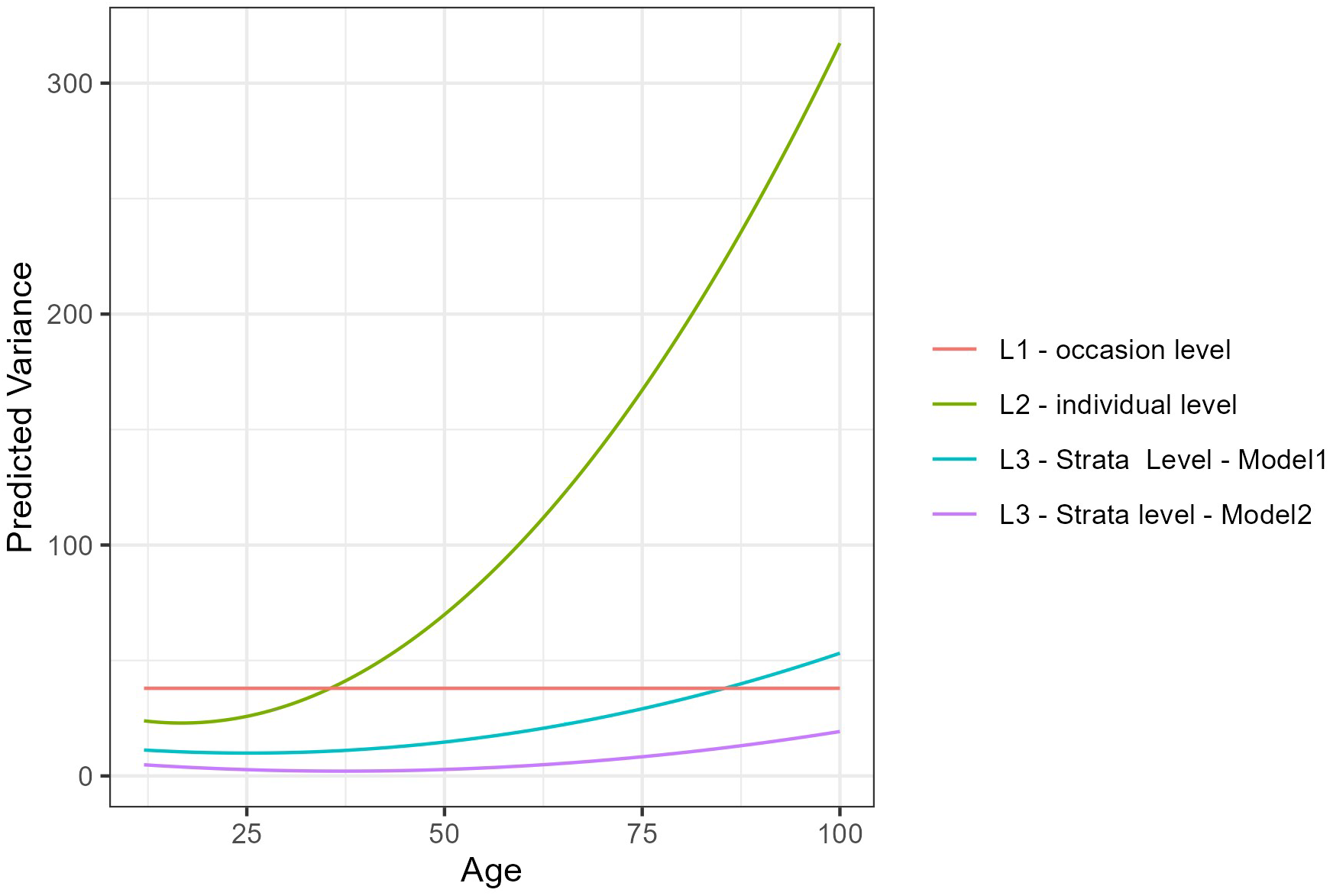
Variance functions plot, SF-12 PCS.

**Figure S2.**
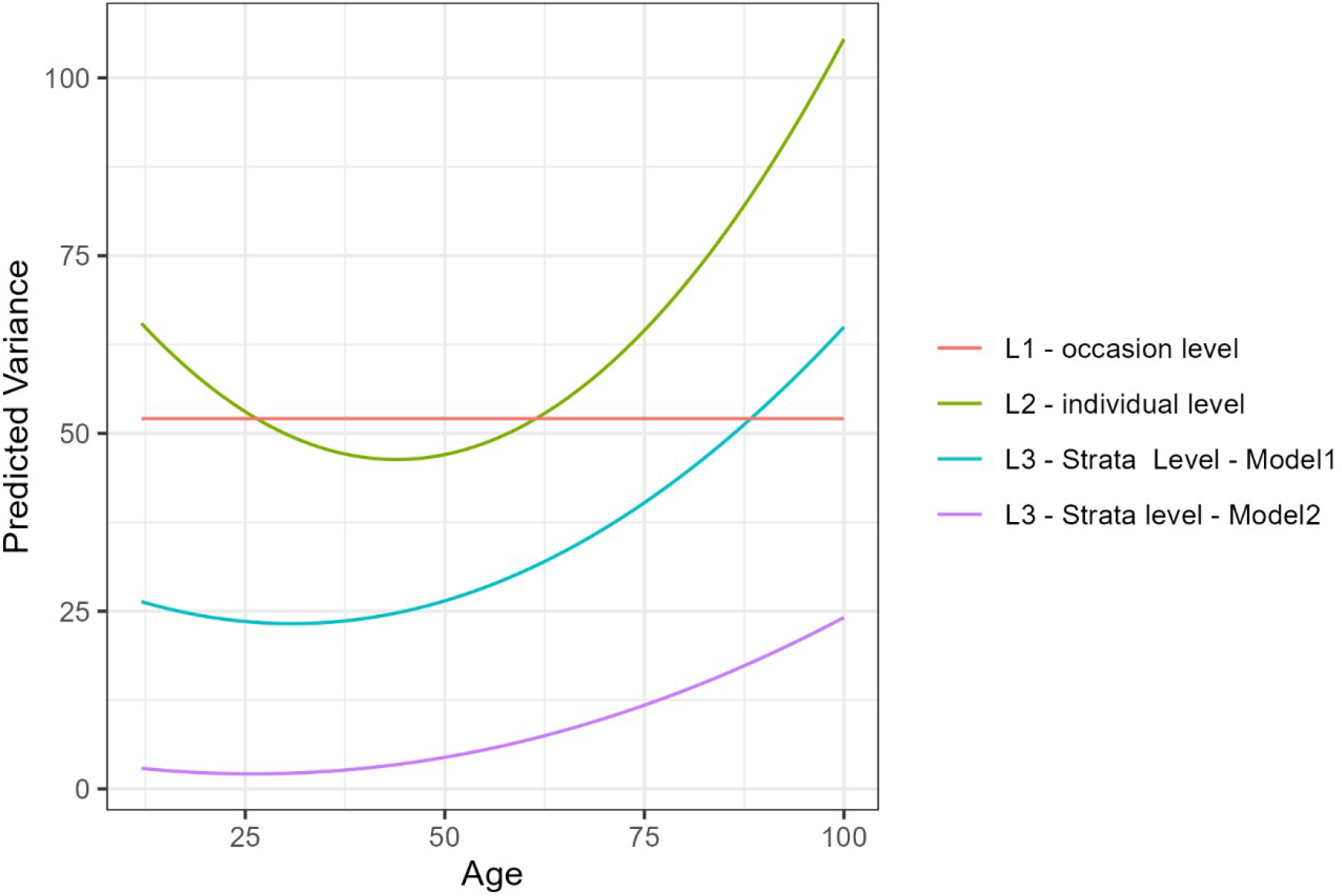
Variance functions plot, SF-12 MCS.

## References

Aboim, S., & Vasconcelos, P. (2014). From political to social generations: A critical reappraisal of Mannheim’s classical approach. European Journal of Social Theory, 17(2), 165–183. 10.1177/1368431013509681

Alberts, S. C., Archie, E. A., Gesquiere, L. R., Altmann, J., Vaupel, J. W., & Christensen, K. (2014). The Male-Female Health-Survival Paradox: A Comparative Perspective on Sex Differences in Aging and Mortality. In Sociality, Hierarchy, Health: Comparative Biodemography: A Collection of Papers. National Academies Press (US). https://www.ncbi.nlm.nih.gov/books/NBK242444/

Arigo, D., Bercovitz, I., Lapitan, E., & Gular, S. (2024). Social Comparison and Mental Health. Current Treatment Options in Psychiatry, 11(2), 17–33. 10.1007/s40501-024-00313-0

Bambra, C., Lynch, J., & Smith, K. E. (2021). *The unequal pandemic: COVID-19 and health inequalities*. Policy press.

Bauer, G. R. (2014). Incorporating intersectionality theory into population health research methodology: Challenges and the potential to advance health equity. Social Science & Medicine, 110, 10–17. 10.1016/j.socscimed.2014.03.022

Bécares, L. (2015). Which ethnic groups have the poorest health? In S. Jivraj & L. Simpson (Eds), Ethnic Identity and Inequalities in Britain: The Dynamics of Diversity (pp. 123–140). Bristol University Press.

Beckfield, J., Olafsdottir, S., & Bakhtiari, E. (2013). Health Inequalities in Global Context. American Behavioral Scientist, 57(8), 1014–1039. 10.1177/0002764213487343

Bell, A. (2014). Life-course and cohort trajectories of mental health in the UK, 1991-2008—A multilevel age-period-cohort analysis. Social Science & Medicine (1982), 120, 21–30. 10.1016/j.socscimed.2014.09.008

Bell, A., Evans, C., Holman, D., & Leckie, G. (2024). Extending intersectional multilevel analysis of individual heterogeneity and discriminatory accuracy (MAIHDA) to study individual longitudinal trajectories, with application to mental health in the UK. Social Science & Medicine, 351, 116955. 10.1016/j.socscimed.2024.116955

Bell, A., Holman, D., & Jones, K. (2019). Using shrinkage in multilevel models to understand intersectionality: A simulation study and a guide for best practice. Methodology.

Ben-Shlomo, Y., Cooper, R., & Kuh, D. (2016). The last two decades of life course epidemiology, and its relevance for research on ageing. International Journal of Epidemiology, 45(4), 973–988. 10.1093/ije/dyw096

Ben-Shlomo, Y., & Kuh, D. (2002). A life course approach to chronic disease epidemiology: Conceptual models, empirical challenges and interdisciplinary perspectives. International Journal of Epidemiology, 31(2), 285–293. 10.1093/ije/31.2.285

Bharadwaj, P., Pai, M. M., & Suziedelyte, A. (2017). Mental health stigma. Economics Letters, 159, 57–60.

Black, M., Adjei, N. K., Strong, M., Barnes, A., Jordan, H., & Taylor-Robinson, D. (2023). Trajectories of Child Cognitive and Socioemotional Development and Associations with Adolescent Health in the UK Millennium Cohort Study. The Journal of Pediatrics, 263, 113611. 10.1016/j.jpeds.2023.113611

Blane, D., Bartley, M., & Smith, G. D. (1997). Disease aetiology and materialist explanations of socioeconomic mortality differentials. The European Journal of Public Health, 7(4), 385–391.

Booker, C. L., Kelly, Y. J., & Sacker, A. (2018). Gender differences in the associations between age trends of social media interaction and well-being among 10-15 year olds in the UK. BMC Public Health, 18(1), 321. 10.1186/s12889-018-5220-4

Bowleg, L. (2012). The Problem With the Phrase *Women and Minorities:* Intersectionality—An Important Theoretical Framework for Public Health. American Journal of Public Health, 102(7), 1267–1273. 10.2105/AJPH.2012.300750

Boyd, A., Van de Velde, S., Vilagut, G., de Graaf, R., O’Neill, S., Florescu, S., Alonso, J., Kovess-Masfety, V., & EU-WMH Investigators. (2015). Gender differences in mental disorders and suicidality in Europe: Results from a large cross-sectional population-based study. Journal of Affective Disorders, 173, 245–254. 10.1016/j.jad.2014.11.002

Burgess, R. A., Chobhthaigh, S. N., Biswal, B., Ceccolini, D., Fadipe, B., Khan, D., Aggarwal, N., Pabla, I., Solis, C., Pillutla, R., Sarkar, P., Frasco, E., lemmi, V., Pathare, S., & Lund, C. (2025). Intersectional discrimination, exclusion and the socio-political economy of global mental health: A systematic scoping review of the literature. SSM - Mental Health, 7, 100382. 10.1016/j.ssmmh.2024.100382

Calasanti, T., & King, N. (2015). Intersectionality and age. Routledge Handbook of Cultural Gerontology, 193–200.

Canetto, S. S., & Sakinofsky, I. (1998). The Gender Paradox in Suicide. Suicide and Life-Threatening Behavior, 28(1), 1–23. 10.1111/j.1943-278X.1998.tb00622.x

Carey, G., Crammond, B., & De Leeuw, E. (2015). Towards health equity: A framework for the application of proportionate universalism. International Journal for Equity in Health, 14(1), 81. 10.1186/s12939-015-0207-6

Case, A., & Deaton, A. (2020). Deaths of despair and the future of capitalism. Princeton university press.

Collins, P. H. (2002). Black Feminist Thought: Knowledge, Consciousness, and the Politics of Empowerment. Routledge. 10.4324/9780203900055

Crawshaw, P., Gray, J., Haighton, C., & Lloyd, S. (2024). Health inequalities and health-related economic inactivity: Why *good* work needs *good* health. Public Health in Practice, 8, 100555. 10.1016/j.puhip.2024.100555

Crenshaw, K. (1989). Demarginalizing the Intersection of Race and Sex: A Black Feminist Critique of Antidiscrimination Doctrine, Feminist Theory and Antiracist Politics. University of Chicago Legal Forum, 139–168.

Crenshaw, K. (1991). Mapping the Margins: Intersectionality, Identity Politics, and Violence against Women of Color. Stanford Law Review, 43(6), 1241–1299. 10.2307/1229039

Crimmins, E. M., Kim, J. K., & Solé-Auró, A. (2011). Gender differences in health: Results from SHARE, ELSA and HRS. European Journal of Public Health, 21(1), 81–91. 10.1093/eurpub/ckq022

Daly, S., & Allen, J. (2016). Inequalities in mental health, cognitive impairment and dementia among older people. UCL Institute of Health Equity.

Dorsett, R., Rienzo, C., & Weale, M. (2019). Intergenerational and interethnic mental health: An analysis for the United Kingdom. Population, Space and Place, 25(2), e2195. 10.1002/psp.2195

Dugravot, A., Fayosse, A., Dumurgier, J., Bouillon, K., Rayana, T. B., Schnitzler, A., Kivimaki, M., Sabia, S., & Singh-Manoux, A. (2019). Social inequalities in multimorbidity, frailty, disability, and transitions to mortality: A 24-year follow-up of the Whitehall II cohort study. *The Lancet*. Public Health, 5(1), e42–e50. 10.1016/S2468-2667(19)30226-9

Elder, G. H., & Giele, J. Z. (2009). Life Course Studies: An Evolving Field. In G. H. Elder & J. Z. Giele (Eds), The Craft of Life Course Research (pp. 1–24). The Guilford Press.

Evandrou, M., Falkingham, J., Feng, Z., & Vlachantoni, A. (2016). Ethnic inequalities in limiting health and self-reported health in later life revisited. Journal of Epidemiology and Community Health, 70(7), 653–662. 10.1136/jech-2015-206074

Evans, C. R., Leckie, G., Subramanian, S. V., Bell, A., & Merlo, J. (2024). A tutorial for conducting intersectional multilevel analysis of individual heterogeneity and discriminatory accuracy (MAIHDA). SSM - Population Health, 26, 101664. 10.1016/j.ssmph.2024.101664

Evans, C. R., Williams, D. R., Onnela, J.-P., & Subramanian, S. V. (2018). A multilevel approach to modeling health inequalities at the intersection of multiple social identities. Social Science & Medicine. 10.1016/j.socscimed.2017.11.011

Fernando, S. (2010). Mental health, race and culture. Bloomsbury Publishing. https://books.google.co.uk/books?hl=en&lr=&id=7h9HEAAAQBAJ&oi=fnd&pg=PR10&dq=Mental+Health,+Race+and+Culture&ots=Vsyo5hCYrc&sig=hGnP-lzHvNAPzQlNCE7rjkZF13w

Ferrer, I., Grenier, A., Brotman, S., & Koehn, S. (2017). Understanding the experiences of racialized older people through an intersectional life course perspective. Journal of Aging Studies, 41, 10–17. 10.1016/j.jaging.2017.02.001

Foulkes, L., & Andrews, J. L. (2023). Are mental health awareness efforts contributing to the rise in reported mental health problems? A call to test the prevalence inflation hypothesis. New Ideas in Psychology, 69, 101010. 10.1016/j.newideapsych.2023.101010

Galenkamp, H., Stronks, K., Mokkink, L. B., & Derks, E. M. (2018). Measurement invariance of the SF-12 among different demographic groups: The HELIUS study. PLOS ONE, 13(9), e0203483. 10.1371/journal.pone.0203483

Gandek, B., Ware, J. E., Aaronson, N. K., Apolone, G., Bjorner, J. B., Brazier, J. E., Bullinger, M., Kaasa, S., Leplege, A., Prieto, L., & Sullivan, M. (1998). Cross-validation of item selection and scoring for the SF-12 Health Survey in nine countries: Results from the IQOLA Project. International Quality of Life Assessment. Journal of Clinical Epidemiology, 51(11), 1171–1178. 10.1016/s0895-4356(98)00109-7

Gilleard, C. (2004). Cohorts and Generations in the Study of Social Change. Social Theory & Health, 2(1), 106–119. 10.1057/palgrave.sth.8700023

Gilmore, A. B., Fabbri, A., Baum, F., Bertscher, A., Bondy, K., Chang, H.-J., Demaio, S., Erzse, A., Freudenberg, N., Friel, S., Hofman, K. J., Johns, P., Karim, S. A., Lacy-Nichols, J., Carvalho, C. M. P. de, Marten, R., McKee, M., Petticrew, M., Robertson, L., … Thow, A. M. (2023). Defining and conceptualising the commercial determinants of health. The Lancet, 401(10383), 1194–1213. 10.1016/S0140-6736(23)00013-2

Gimeno, L., Goisis, A., Dowd, J. B., & Ploubidis, G. B. (2024). Cohort Differences in Physical Health and Disability in the United States and Europe. The Journals of Gerontology: Series B, 79(8), gbae113. 10.1093/geronb/gbae113

Gimeno, L., Moreno-Agostino, D., Danka, M., Guo, Y., Goisis, A., Dowd, J. B., & Ploubidis, G. B. (2025). The Generational Health Drift: A Systematic Review of Evidence from the British Birth Cohort Studies. Epidemiology. 10.1101/2025.06.12.25329414

Guan, S., Coughlan, B., Evans, K., & Duschinsky, R. (2024). Associations between ethnicity and mental health problems among children and adolescents in the United Kingdom: A systematic review and narrative synthesis. BMC Public Health, 24(1), 3267. 10.1186/s12889-024-20695-3

Hayanga, B., Stafford, M., & Bécares, L. (2023). Ethnic inequalities in multiple long-term health conditions in the United Kingdom: A systematic review and narrative synthesis. BMC Public Health, 23(1), 178. 10.1186/s12889-022-14940-w

Hesketh, K. R., Law, C., Bedford, H., & Hope, S. (2016). Co-Occurrence of Health Conditions during Childhood: Longitudinal Findings from the UK Millennium Cohort Study (MCS). PLOS ONE, 11(6), e0156868. 10.1371/journal.pone.0156868

Holman, D., & Walker, A. (2020). Understanding unequal ageing: Towards a synthesis of intersectionality and life course analyses. European Journal of Ageing. 10.1007/s10433-020-00582-7

Homan, P., Brown, T. H., & King, B. (2021). Structural Intersectionality as a New Direction for Health Disparities Research. Journal of Health and Social Behavior, 62(3), 350–370. 10.1177/00221465211032947

Hossin, M. Z., Björk, J., & Koupil, I. (2020). Early-life social and health determinants of adult socioeconomic position: Associations and trends across generations. J Epidemiol Community Health, 74(5), 412–420. 10.1136/jech-2019-213209

Houweling, T. A. J., & Grünberger, I. (2024). Intergenerational transmission of health inequalities: Towards a life course approach to socioeconomic inequalities in health – a review. J Epidemiol Community Health, 78(10), 641–649. 10.1136/jech-2022-220162

Jones, N. L., Gilman, S. E., Cheng, T. L., Drury, S. S., Hill, C. V., & Geronimus, A. T. (2019). Life Course Approaches to the Causes of Health Disparities. American Journal of Public Health, 109(Suppl 1), S48–S55. 10.2105/AJPH.2018.304738

Kapadia, D., Zhang, J., Salway, S., Nazroo, J., Booth, A., Villarroel-Williams, N., Bécares, L., & Esmail, A. (2022). Ethnic Inequalities in Healthcare: A Rapid Evidence Review. NHS Race & Health Observatory. https://www.nhsrho.org/wp-content/uploads/2022/02/RHO-Rapid-Review-Final-Report_v.7.pdf

Khanolkar, A. R., Chaturvedi, N., Kuan, V., Davis, D., Hughes, A., Richards, M., Bann, D., & Patalay, P. (2021). Socioeconomic inequalities in prevalence and development of multimorbidity across adulthood: A longitudinal analysis of the MRC 1946 National Survey of Health and Development in the UK. PLOS Medicine, 18(9), e1003775. 10.1371/journal.pmed.1003775

Konstantinou, G., & Attia, M. (2023). Perspective Chapter: From the Boom to Gen Z – Has Depression Changed across Generations? In F. Durbano, F. Irtelli, & B. Marchesi (Eds), Depression—What Is New and What Is Old in Human Existence. IntechOpen. 10.5772/intechopen.1003091

Kuh, D., Susser, E., Blodgett, J. M., & Ben-Shlomo, Y. (2025). *A Life Course Approach to the Epidemiology of Chronic Diseases and Ageing*. Oxford University Press. https://books.google.co.uk/books?hl=en&lr=&id=GBlAEQAAQBAJ&oi=fnd&pg=PP1&dq=A+Life+Course+Approach+to+the+Epidemiology+of+Chronic+Diseases+and+Ageing+(A+Life+Course+Approach+to+Adult+Health)+kuh+2025&ots=HPEiOFmf2U&sig=JCZPaWBu37Q-SHl-gzLfB4hZnfk

Leckie, G., Bell, A., Merlo, J., Subramanian, S., & Evans, C. (2025). The Statistical Advantages of Multilevel Analysis of Individual Heterogeneity and Discriminatory Accuracy for Estimating Intersectional Inequalities. Sociological Methods & Research, 00491241251385123. 10.1177/00491241251385123

Link, B. G., & Phelan, J. (1995). Social Conditions As Fundamental Causes of Disease. Journal of Health and Social Behavior, 35, 80–94. 10.2307/2626958

Marmot, M., Allen, J., Boyce, T., Goldblatt, P., & Morrison, J. (2020). Health equity in England: The Marmot Review 10 years on. Institute of Health Equity.

McCall, L. (2005). The Complexity of Intersectionality. Signs, 30(3), 1771–1800. 10.1086/426800

McDonough, P., Worts, D., Booker, C., McMunn, A., & Sacker, A. (2015). Cumulative disadvantage, employment–marriage, and health inequalities among American and British mothers. Advances in Life Course Research, 25, 49–66. 10.1016/j.alcr.2015.05.004

McFall, S., Nandi, A., & Platt, L. (2024). *Understanding society: UK household longitudinal study: user guide to ethnicity and immigration research*. 12th Edition. University of Essex.

McGorry, P., Gunasiri, H., Mei, C., Rice, S., & Gao, C. X. (2025). The youth mental health crisis: Analysis and solutions. Frontiers in Psychiatry, 15. 10.3389/fpsyt.2024.1517533

Nazroo, J. Y. (2003). The Structuring of Ethnic Inequalities in Health: Economic Position, Racial Discrimination, and Racism. American Journal of Public Health, 93(2), 277–284.

NHS England. (2025). Adult Psychiatric Morbidity Survey: Survey of Mental Health and Wellbeing, England. https://digital.nhs.uk/data-and-information/publications/statistical/adult-psychiatric-morbidity-survey/survey-of-mental-health-and-wellbeing-england-2023-24/common-mental-health-conditions

Office for National Statistics. (2021). Ethnic differences in life expectancy and mortality from selected causes in England and Wales: 2011 to 2014. Office for National Statistics. https://backup.ons.gov.uk/wp-content/uploads/sites/3/2021/07/Ethnic-differences-in-life-expectancy-and-mortality-from-selected-causes-in-England-and-Wales-2011-to-2014.pdf

Parra-Mujica, F., Robson, M., & Cookson, R. (2021). Socioeconomic Health Inequalities: Differences Between and Within Individuals. HEDG, c/o Department of Economics, University of York. https://ideas.repec.org/p/yor/hectdg/21-15.html

Pearce, A., Dundas, R., Whitehead, M., & Taylor-Robinson, D. (2019). Pathways to inequalities in child health. Archives of Disease in Childhood, 104(10), 998–1003. 10.1136/archdischild-2018-314808

Petrovic, D., de Mestral, C., Bochud, M., Bartley, M., Kivimäki, M., Vineis, P., Mackenbach, J., & Stringhini, S. (2018). The contribution of health behaviors to socioeconomic inequalities in health: A systematic review. Preventive Medicine, 113, 15–31. 10.1016/j.ypmed.2018.05.003

Pickett, K., & Wilkinson, R. (2009). The Spirit Level: Why more equal societies almost always do better. London: Allen Lane.

Reed, H. R., Nettle, D., Parra-Mujica, F., Stark, G., Wilkinson, R., Johnson, M. T., & Johnson, E. A. (2025). Examining the relationship between income and both mental and physical health among adults in the UK: Analysis of 12 waves (2009-2022) of Understanding Society. PloS One, 20(3), e0316792. 10.1371/journal.pone.0316792

Richardson, L. J., & Brown, T. H. (2016). (En)gendering Racial Disparities in Health Trajectories: A Life Course and Intersectional Analysis. SSM - Population Health, 2, 425–435. 10.1016/j.ssmph.2016.04.011

Riecher-Rössler, A. (2017). Sex and gender differences in mental disorders. The Lancet Psychiatry, 4(1), 8–9. 10.1016/S2215-0366(16)30348-0

Robertson, R., Williams, E., Buck, D., & Breckwoldt, J. (2021). Ethnic health inequalities and the NHS. The King’s Fund, Commissioned by the NHS Race and Health Observatory. https://healthwatchgreenwich.co.uk/sites/healthwatchgreenwich.co.uk/files/Ethnic-Health-Inequalities-Kings-Fund-Report.pdf

Rose, D., & Pevalin, D. J. (2003). A researcher’s guide to the national statistics socio-economic classification. SAGE Publications Ltd.

Rudolph, C. W., & Zacher, H. (2022). Generations, We Hardly Knew Ye: An Obituary. Group & Organization Management, 47(5), 928–935. 10.1177/10596011221098307

Stopforth, S., Kapadia, D., Nazroo, J., & Bécares, L. (2021). Ethnic inequalities in health in later life, 1993–2017: The persistence of health disadvantage over more than two decades. Ageing & Society, 1–29. 10.1017/S0144686X2100146X

Thoits, P. A. (2013). Self, Identity, Stress, and Mental Health. In C. S. Aneshensel, J. C. Phelan, & A. Bierman (Eds), Handbook of the Sociology of Mental Health (pp. 357–377). Springer Netherlands. 10.1007/978-94-007-4276-5_18

Vlassoff, C. (2007). Gender Differences in Determinants and Consequences of Health and Illness. Journal of Health, Population, and Nutrition, 25(1), 47–61.

Wallace, S., Nazroo, J., & Bécares, L. (2016). Cumulative Effect of Racial Discrimination on the Mental Health of Ethnic Minorities in the United Kingdom. American Journal of Public Health, 106(7), 1294–1300. 10.2105/AJPH.2016.303121

Ware, J. E., Kosinski, M., & Keller, S. D. (1995). SF-12: How to Score the SF-12 Physical and Mental Health Summary Scales (Second Edition). The Health Institute, New England Medical Center.

Ware, J. E., Kosinski, M., & Keller, S. D. (1996). A 12-Item Short-Form Health Survey: Construction of Scales and Preliminary Tests of Reliability and Validity. Medical Care, 34(3), 220.

Williams, D. R., Lawrence, J. A., & Davis, B. A. (2019). Racism and Health: Evidence and Needed Research. Annual Review of Public Health, 40, 105–125. 10.1146/annurev-publhealth-040218-043750

Williams, E. D., Cox, A., & Cooper, R. (2020). Ethnic Differences in Functional Limitations by Age Across the Adult Life Course. The Journals of Gerontology Series A: Biological Sciences and Medical Sciences, 75(5), 914–921. 10.1093/gerona/glz264

Zaninotto, P., Batty, G. D., Stenholm, S., Kawachi, I., Hyde, M., Goldberg, M., Westerlund, H., Vahtera, J., & Head, J. (2020). Socioeconomic Inequalities in Disability-free Life Expectancy in Older People from England and the United States: A Cross-national Population-Based Study. The Journals of Gerontology Series A: Biological Sciences and Medical Sciences, 75(5), 906–913. 10.1093/gerona/glz266

